# Evaluation of ComBat harmonization for reducing across-tracer differences in regional amyloid PET analyses

**DOI:** 10.1101/2024.06.14.24308952

**Authors:** Braden Yang, Tom Earnest, Sayantan Kumar, Deydeep Kothapalli, Tammie Benzinger, Brian Gordon, Aristeidis Sotiras

## Abstract

**Introduction:** Differences in amyloid positron emission tomography (PET) radiotracer pharmacokinetics and binding properties lead to discrepancies in amyloid-β uptake estimates. Harmonization of tracer-specific biases is crucial for optimal performance of downstream tasks. Here, we investigated the efficacy of ComBat, a data-driven harmonization model, for reducing tracer-specific biases in regional amyloid PET measurements from [^18^F]-florbetapir (FBP) and [^11^C]-Pittsburgh Compound-B (PiB).

**Methods:** One-hundred-thirteen head-to-head FBP-PiB scan pairs, scanned from the same subject within ninety days, were selected from the Open Access Series of Imaging Studies 3 (OASIS-3) dataset. The Centiloid scale, ComBat with no covariates, ComBat with biological covariates, and GAM-ComBat with biological covariates were used to harmonize both global and regional amyloid standardized uptake value ratios (SUVR). Variants of ComBat, including longitudinal ComBat and PEACE, were also tested. Intraclass correlation coefficient (ICC) and mean absolute error (MAE) were computed to measure the absolute agreement between tracers. Additionally, longitudinal amyloid SUVRs from an anti-amyloid drug trial were simulated using linear mixed effects modeling. Differences in rates-of-change between simulated treatment and placebo groups were tested, and change in statistical power/Type-I error after harmonization was quantified.

**Results:** In the head-to-head tracer comparison, ComBat with no covariates was the best at increasing ICC and decreasing MAE of both global summary and regional amyloid PET SUVRs between scan pairs of the same group of subjects. In the clinical trial simulation, harmonization with both Centiloid and ComBat increased statistical power of detecting true rate-of-change differences between groups and decreased false discovery rate in the absence of a treatment effect. The greatest benefit of harmonization was observed when groups exhibited differing FBP-to-PiB proportions.

**Conclusion:** ComBat outperformed the Centiloid scale in harmonizing both global and regional amyloid estimates. Additionally, ComBat improved the detection of rate-of-change differences between clinical trial groups. Our findings suggest that ComBat is a viable alternative to Centiloid for harmonizing regional amyloid PET analyses.

## Introduction

Positron emission tomography (PET) is widely used in clinical and research settings for measuring and monitoring amyloid-β deposition *in vivo* in the brain for patients who are at risk of developing or who already present with Alzheimer’s disease (AD). In clinical trials for anti-amyloid drugs, PET is an important tool for screening appropriate candidates who have undergone significant amyloidosis in the brain [1]. Moreover, PET has also been used for monitoring the progression of global amyloid burden longitudinally within these trials, which along with measures of cognitive function serves as a crucial secondary endpoint [2,3]. In research settings, PET is able to resolve the spatial distribution of amyloid within specific regions of the brain, enabling the design of multivariable statistical analyses and predictive models of AD using voxel-wise [4,5] or region-of-interest (ROI) based [6–8] PET biomarkers as multidimensional features.

Several PET radiotracers for imaging brain amyloid pathology have been developed. The first amyloid PET tracer developed for human imaging studies was [^11^C]-Pittsburgh compound B (PiB) [9], but due to its short half-life requires an on-site cyclotron to produce. Consequently, PiB is not accessible by many sites and not appropriate for use in clinical trials. Alternatively, amyloid measurements obtained from ^18^F-based tracers such as [^18^F]-florbetapir (FBP) [10–12], [^18^F]-florbetaben [13] and [^18^F]-flutemetamol [14,15] have been shown to correlate well with PiB. Coupled with a much longer half-life than PiB, these tracers are a much more suitable option for clinical trials due to their accessibility and ability to be distributed off-site.

Nonetheless, previous studies that performed a head-to-head comparison of amyloid PET tracers have demonstrated significant disparities in dynamic range and non-specific binding properties between tracers [10,13,16]. Subsequently, this makes it difficult to compare quantitative amyloid measurements between images acquired using different tracers. This may also negatively impact the performance of downstream tasks such as detecting significant treatment effects in anti-amyloid drug trials [17].

To address this, Klunk *et al.* introduced the Centiloid scale [18], which linearly transforms the dynamic range of a global estimate of amyloid burden to a common scale and converts it to Centiloid (CL) units. This involves calibrating the scale to a preselected cohort of amyloid-negative healthy controls and amyloid-positive typical AD patients, where the average global burden of the two groups are set to 0 CL and 100 CL, respectively. However, the calibration process requires at least two PET scans from the same subject within a short time period in order to calibrate conversion equations. Additionally, a single equation is usually derived to operate on the global amyloid estimate, but this cannot address local disparities in amyloid PET signal between tracers. Other methods for tracer harmonization that are based on data-driven and/or machine learning techniques such as principal component analysis [19], non-negative matrix factorization [20], and deep learning [21,22] have been proposed, but like Centiloid they focus on the global amyloid burden.

Alternatively, ComBat [23] is a data-driven harmonization model which has been widely applied in magnetic resonance imaging (MRI) analyses to adjust for differences in scanners and acquisition protocols. It has been used to correct regional volume and cortical thickness measurements from MRI [24–27], and has more recently been applied to [^18^F]-fluorodeoxyglucose-PET [28] and amyloid PET [29] biomarkers. Much of the current literature on applying ComBat has focused on reducing scanner-level and institutional-level biases. However, it remains unclear whether ComBat is applicable for mitigating across-tracer variance, specifically in regional amyloid PET measurements.

Here, we aimed to evaluate the efficacy of ComBat for harmonizing standardized uptake value ratios (SUVR) from amyloid PET across two tracers - PiB and FBP. Specifically, we addressed two primary inquiries. Firstly, we investigated whether ComBat harmonization may increase the agreement between regional SUVRs obtained from the two tracers. This was accomplished through a head-to-head comparison of PiB and FBP. We selected a set of PiB-FBP scan pairs acquired from the same subject in a short time period and compared measures of the absolute agreement between regional SUVRs before and after ComBat harmonization. Secondly, we explored the utility of ComBat harmonization in the context of clinical tasks. This was examined by simulating a multi-tracer anti-amyloid drug trial where two different amyloid tracers were used to measure brain amyloid deposition, under the assumption that different sites have access to different tracers. We generated longitudinal amyloid PET data of hypothetical treatment and placebo groups with a known underlying treatment effect, and assigned each group a specific proportion of PiB-to-FBP scans. We then gauged whether ComBat harmonization improves the statistical power of detecting the underlying treatment effect when using two different tracers.

## Materials & Methods

### Participants and data

Data for this study were acquired from the Open Access Series of Imaging Studies 3 (OASIS-3) dataset [30], which consisted of 1098 total participants and their longitudinal imaging data. Of these, we selected 997 who underwent PiB and/or FBP amyloid PET imaging. All available PET scans, including the initial baseline scan and any follow-up scans, were utilized in this study, for a total of 678 FBP scans and 1157 PiB scans. Additionally, each subject’s age at scan, sex and apolipoprotein-ε4 (APOE) allele carriership were extracted. Subjects who were missing any of these variables were excluded from further analyses.

### Image acquisition and processing

All amyloid PET imaging from OASIS-3 were acquired at Washington University in St. Louis using one of four Siemens scanner models: Biograph mMR PET/MR 3T, Biograph 40 PET/CT, Biograph 128 Vision Edge PET/CT, and ECAT HR+ 962 PET. For PiB PET, participants received a bolus injection of 6-20 mCi of PiB, and a 60-minute dynamic scan was acquired. For FBP PET, participants received a bolus injection of 10 mCi of FBP, and either a 70-minute dynamic scan was acquired, or a 20-minute dynamic scan was acquired at 50-minutes post-injection. Additionally, T1-weighted MRI scans were acquired and utilized for PET processing. All MRI imaging from the OASIS-3 dataset were acquired at Washington University in St. Louis using one of three Siemens scanner models: Vision 1.5T, TIM Trio 3T, and Biograph mMR PET/MR 3T.

PET images were processed using the PET Unified Pipeline (https://github.com/ysu001/PUP), described in [31]. Briefly, raw PET images were smoothed to 8mm spatial resolution, corrected for inter-frame motion, and coregistered to the T1 MRI scan acquired closest in time using a vector-gradient algorithm. T1 images were segmented and parcellated into cortical and subcortical ROIs using FreeSurfer 5.0 or 5.1 for 1.5T scans or FreeSurfer 5.3 for 3T scans. For each ROI, regional SUVRs were computed from the peak time windows of each tracer (30-to-60 minutes post-injection for PiB, 50-to-70 minutes post-injection for FBP). The average of the left and right cerebellar cortex was used as the reference region. Additionally, a summary estimate of global amyloid burden was derived by computing the SUVR of a meta-ROI comprised of lateral and medial orbitofrontal, middle and superior temporal, superior frontal, rostral middle frontal, and precuneus ROIs from both hemispheres. For subsequent analyses, we chose to focus on 68 cortical, 16 subcortical regions, and the global summary region. The full list of regions is given in Supp. Table S1.

### Data harmonization

Five harmonization methods were investigated in the current study: Centiloid [18], ComBat [23,27], GAM-ComBat [26], longitudinal ComBat [32], and Probabilistic Estimation for Across-batch Compatibility Enhancement (PEACE) [29]. These methods are briefly described below.

### Centiloid

The Centiloid scale [18] is a method of linearly transforming global amyloid burden estimates from SUVRs to a scale that is standardized across tracers. Centiloid ranges from 0 to 100, with 0 corresponding to the average amyloid burden of a group of healthy controls, and 100 corresponding to the average amyloid burden of typical AD patients. Note that Centiloids are allowed to fall above 100 CL or below 0 CL.

Although Centiloid is calibrated against and primarily used to harmonize the global summary SUVR, it can also be applied to regional or voxel-wise SUVRs [17,18]. To convert regional SUVRs to Centiloid, we utilized the conversion equations that were previously validated for the OASIS-3 cohort [10,33]:

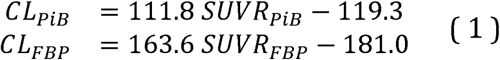

### ComBat

ComBat is a data-driven method for adjusting data with batch-specific effects [23], where batches refer to any nominal variable(s) which may contribute confounding biases in the target measurement. It utilizes a multivariable linear regression to model measurements in terms of batch-specific shift and scale parameters, as well as other covariates which model variance due to biologically relevant effects. For batch effect *i*;, subject *j* and feature *k*, ComBat models the measurement, *y_ijk_* as:

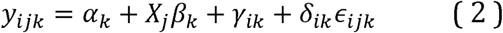

where α_k_ is the mean measurement across all subjects and all batches, *X_j_* is the vector of biological covariates associated with subject *j*, and β*_k_* is the vector of coefficients for *X_j_*. The batch-specific shift (additive) and scale (multiplicative) parameters are represented by γ*_ik_* and δ*_ik_* respectively. These modify the measurement from the group average to account for batch-specific biases. ϵ,_*ijk*_ is the error term, which is assumed to be normally distributed with zero mean and unit variance. γ_*ik*_ and δ_*ik*_ are estimated using an empirical Bayesian approach, and once estimated, the measurement without batch effects can be recovered by the following:

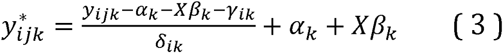

This adjustment ensures that only variance due to the batch effects is corrected for, while variance due to the covariates is preserved, which is a unique advantage of ComBat over other batch-adjusting techniques. In subsequent experiments, we selected age, sex, and APOE carriership as the covariates of interest to preserve.

### GAM-ComBat

A limitation of the ComBat model is that it is only able to model covariates as linearly related to the target variable. To address this, Pomponio *et al.* [26] developed GAM-ComBat, a variant of ComBat which can model continuous covariates non-linearly using generalized additive models (GAM):

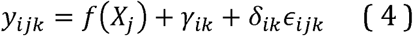

where *f* is the GAM. In subsequent experiments, we explored modeling the age covariate non-linearly using GAMs.

### Other ComBat variants

We also tested two variants of ComBat: longitudinal ComBat [32] and PEACE [29]. Longitudinal ComBat incorporates a subject-specific random intercept term to the original ComBat model:

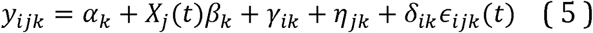

where η*_jk_* is the subject-specific random intercept, which is preserved after harmonization. Additionally, the covariate design matrix *X_j_* and error term η*_ijk_* are parameterized by time *t*, allowing for these terms to be varied across time. This model is appropriate for harmonizing data consisting of multiple repeated measurements of the same subjects at different time points.

PEACE differs from ComBat in two aspects: (1) it models target measurements using a bimodal Gaussian mixture model to estimate two clusters of the data, then estimates the batch-specific parameters independently of the cluster assignments; and (2) rather than estimating model parameters and hyperparameters in an empirical Bayesian manner, it employs a fully Bayesian approach where these parameters are assumed to be distributed by fixed priors. PEACE addresses ComBat’s limitation of assuming that the target features, after residualizing covariate terms, are distributed normally. However, this is often not the case for amyloid imaging data, where the distribution of amyloid burden across subjects often exhibits a bimodal pattern of low amyloid (amyloid-negative) and high amyloid (amyloid-positive) clusters [29,34].

The PEACE model is given by the following:

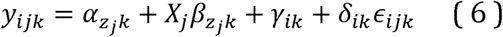

where *z_j_* indicates the cluster assignment of subject *j*. PEACE is fit using Hamiltonian Monte Carlo Markov chain sampling. In our experiments, we ran 4 separate Markov chains where 50 warmup iterations were sampled followed by 50 samples drawn from the posterior, for a total of 200 samples after warmup. We then averaged over these 200 samples to obtain the final harmonized data.

The original implementation of PEACE only considered a single covariate. We modified the model to allow for either no covariates or multiple covariates. In subsequent experiments, we selected age, sex, and APOE carriership as the covariates of interest to preserve. We further configured PEACE to train on one dataset and be applied on held-out data.

### Statistical analysis

To evaluate Centiloid and ComBat for harmonizing inter-tracer differences for both global and regional amyloid PET features, we performed two experiments: a head-to-head comparison of FBP and PiB to evaluate absolute agreement, and a clinical trial simulation to evaluate the clinical utility of harmonization. A summary of the pipelines for the two experiments and the data used for each is illustrated in Fig. 1.

**Fig. 1.**
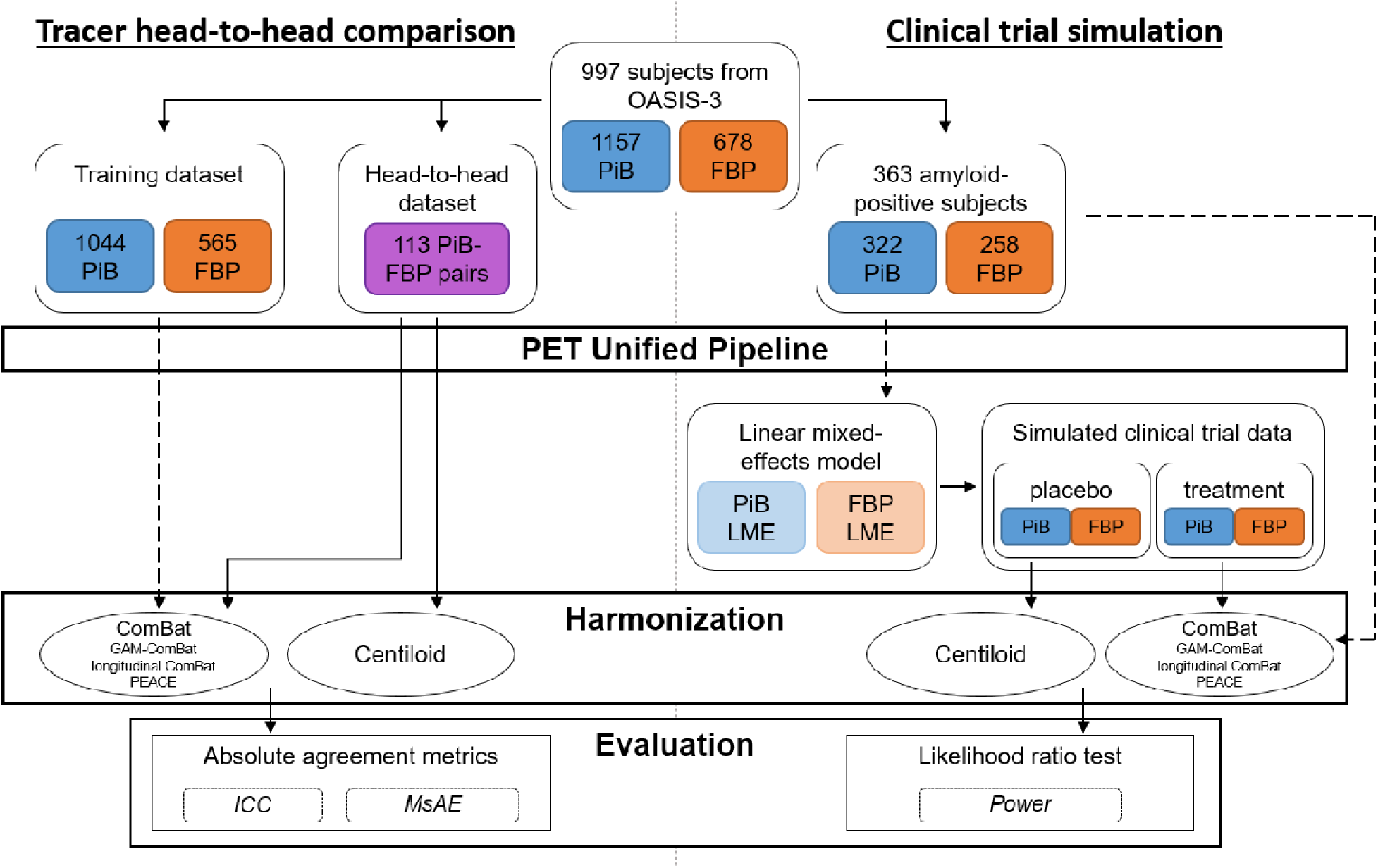
Flowchart of data for the tracer head-to-head comparison (left) and clinical trial simulation (right). Dotted arrows indicate where data was used to train ComBat or linear mixed effects models.

### Tracer head-to-head comparison

We performed a head-to-head comparison of FBP and PiB measurements and evaluated their absolute agreement after harmonization. We identified 113 FBP-PiB scan pairs across 99 subjects which were acquired within 90 days. All remaining scans were used to train ComBat models, which included 565 FBP and 1044 PiB scans.

Centiloid, three different configurations of ComBat, and PEACE were applied to the global summary SUVR and 84 regional SUVRs from the head-to-head dataset to harmonize tracer differences. We tested ComBat without any covariates, ComBat with age, sex and APOE-,4 carriership as linear covariates, and GAM-ComBat with sex and APOE-4 carriership as linear covariates and age as a non-linear covariate. Additionally, we tested PEACE without any covariates and PEACE with age, sex and APOE-4 carriership as linear covariates. Note that we omitted longitudinal ComBat from this analysis since the head-to-head data was treated as cross-sectional.

To evaluate the absolute agreement between FBP and PiB measurements, two metrics were computed. Firstly, intraclass correlation coefficient (ICC) using a fixed rater, single measurement model (i.e., ICC3) was estimated. ICC is roughly the ratio of intraclass variance to total variance, and values closer to 1 indicate better agreement between the two tracers. Secondly, the absolute error (AE) between FBP and PiB measurements was computed:

where *PiB_i_* and *FBP_i_* are the measurements made with PiB and FBP from scan pair *i*. We also computed the mean absolute error (MAE) across all scan pairs:

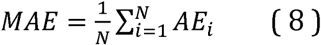

where *N* is the number of scan pairs. To facilitate comparisons of absolute errors between (un)harmonized SUVRs and Centiloids, we scaled Centiloids to a similar dynamic range as SUVRs. Utilizing the Centiloid conversion equations in Equation 1, we computed the *SUVR_FBP_* and *SUVR_PiB_* that result in 0 CL and 100 CL (denoted as *SUVR*_{*tracer*},0*CL*_ and *SUVR*_{*tracer*},100*CL*_, respectively). Then, using the average of FBP and PiB SUVRs at these anchor points, we linearly mapped Centiloids back to SUVR using the following equation:

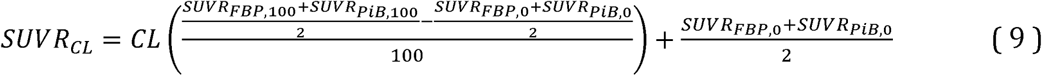

Substituting the respective SUVRs (*SUVR*_*FBP*,100_ = 1.718, *SUVR*_*PiB*,100_ = 1.961, *SUVR*_*FBP*,0_ = 1.106, *SUVR*_*PiB*,0_ = 1.067) into the above equation yields the following:

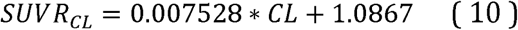

We will refer to this as the scaled Centiloid in the remainder of the text.

Paired t-tests were performed to test for significant differences in the distributions of ICC and MAE between unharmonized SUVRs and each of the four harmonization methods. Additionally, we further subdivided each FreeSurfer region into three groups – regions belonging to the global summary meta-ROI, other cortical regions not part of the summary meta-ROI, and subcortical regions. We then performed paired t-tests for each group separately to compare across harmonization methods. In all statistical tests, Bonferroni correction was applied to correct for multiple comparisons.

### Clinical trial simulation

We evaluated Centiloid and ComBat in the context of improving detection of treatment effects in an anti-amyloid drug trial setting, with the assumption that multiple amyloid PET tracers were used due to pooling of data from multiple institutions. To accomplish this, we modeled a simulation experiment after those described in Chen *et al.* [17] to generate data of placebo and treatment groups. We varied the proportion of FBP-to-PiB scans of each group, then tested for group differences of amyloid rate-of-change.

We selected subjects who presented as PET amyloid-positive at least once during their participation in OASIS-3. To mark scans as amyloid-positive, we used a global summary SUVR threshold of 1.31 for PiB and 1.24 for FBP. These thresholds were previously validated for the OASIS-3 cohort [10]. From these criteria, we identified 363 amyloid-positive subjects, from which 258 FBP and 322 PiB scans were selected.

For each tracer and for each region-of-interest (including the global summary region), a linear mixed effects (LME) model was fit on the selected scans to predict longitudinal SUVR. Sex, APOE carriership, baseline age, and time-from-baseline were specified as fixed effects. A random intercept grouped by subject was specified as the only random effect. This resulted in two LME models – one fitted to FBP data and one to PiB data – for each region-of-interest. Fitted LME models were then used to generate new longitudinal data of placebo and treatment groups. For the placebo group, the models were applied as is to generate SUVRs that follow the natural longitudinal trajectory among amyloid-positive subjects in OASIS-3. For the treatment group, we added a negative rate-of-change term to the LME equation to mimic a treatment effect. We tested multiple values of the treatment effect from 0 to -0.03 SUVR, varying in increments of -0.01 SUVR. These values were chosen based off of previously reported clinical trial effect sizes [2,17].

To simulate a single subject’s data, we randomly sampled the empirical distributions of the number of longitudinal scans, age at baseline scan, and interval between scans among the OASIS-3 amyloid-positive cohort to generate longitudinal time points. Each simulated subject was randomly assigned sex and APOE carriership based on the empirical distributions of these covariates. We then allocated a tracer (either PiB or FBP) to each time point, with the following constraints: (1) the proportion of tracers across all scans from all subjects approximated a prespecified proportion; (2) a subject could only switch tracers once during their clinical trial participation, reflecting a realistic scenario where multiple tracers are utilized in a single study. Time points assigned to PiB or FBP were then input into the corresponding trained LME model to obtain simulated SUVR measurements. For our experiments, we varied the percentage of FBP scans from 0.1 to 0.9 in increments of 0.2 for both clinical trial groups independently. We fixed the total number of subjects to 50 per group, with the number of scans per subject ranging from 2 to 6, the mean (± standard deviation) time in between scans being 3.35 ± 1.46 years, and the mean (± standard deviation) baseline age being 69.4 ± 9.2 years.

The simulated data was harmonized using one of four methods: Centiloid, ComBat, PEACE, or longitudinal ComBat. We tested ComBat, PEACE and longitudinal ComBat both without covariates and with age, sex, and APOE-ε4 carriership as linear covariates. Note that we omitted GAM-ComBat from this analysis, since the simulated data was generated using a linear age term in the LME. ComBat and PEACE were trained using all available amyloid-positive scans. For longitudinal ComBat, since the random intercept terms are indexed by subjects in the training dataset, it is not possible to use this model to harmonize data for new subjects. Therefore, we trained longitudinal ComBat on the simulated subjects’ data, and trained models were applied to harmonize this data.

To test for group differences in the rate-of-change in amyloid SUVR between placebo and treatment groups, we first fitted the same LME described previously, but with three additional terms – clinical trial group, interaction of time-from-baseline with trial group, and tracer. We then tested for statistical significance of the time-from-baseline and clinical trial group interaction term, which would indicate whether the two groups exhibit different rates-of-change. Likelihood ratio tests were used to compare the fit of the full model with a nested model that excludes this term, and significance was determined using α = 0.05. A one-way test was used, meaning that we considered a rate-of-change difference to be statistically significant only if the treatment group had a lower rate-of-change than the placebo. Simulations were repeated 1000 times for each permutation of tracer mixing proportions and treatment effect. Statistical power was computed as the proportion of simulation iterations which resulted in a significant finding. Note that for a treatment effect of zero, i.e. the absence of a ground truth treatment effect, this corresponds to the Type-I error rate.

## Results

### Demographics

Descriptive statistics of each cohort are listed in Table 1. A two-tailed t-test was used to test for differences in age at scan, and Fisher’s exact test was used to test for differences in sex, APOE-ε4, and Clinical Dementia Rating^©^ (CDR). Significant differences in age and CDR were observed between the head-to-head cohort and the single-tracer FBP cohort (p < 0.005). Age was also significantly different between the head-to-head and the mixed-tracer PiB cohorts (p < 0.05), and CDR was significantly different between the head-to-head and single-tracer PiB cohorts (p < 0.005). For the tracer-vs-tracer comparison, age was significantly different in both single- and mixed-tracer cohorts in the training dataset (p < 0.005), and for only the mixed-tracer cohort in the simulation dataset (p < 0.005). Lastly, sex and APOE-ε4 carriership were significantly different between FBP and PiB in the single-tracer simulation dataset (p < 0.01).

**Table 1.**
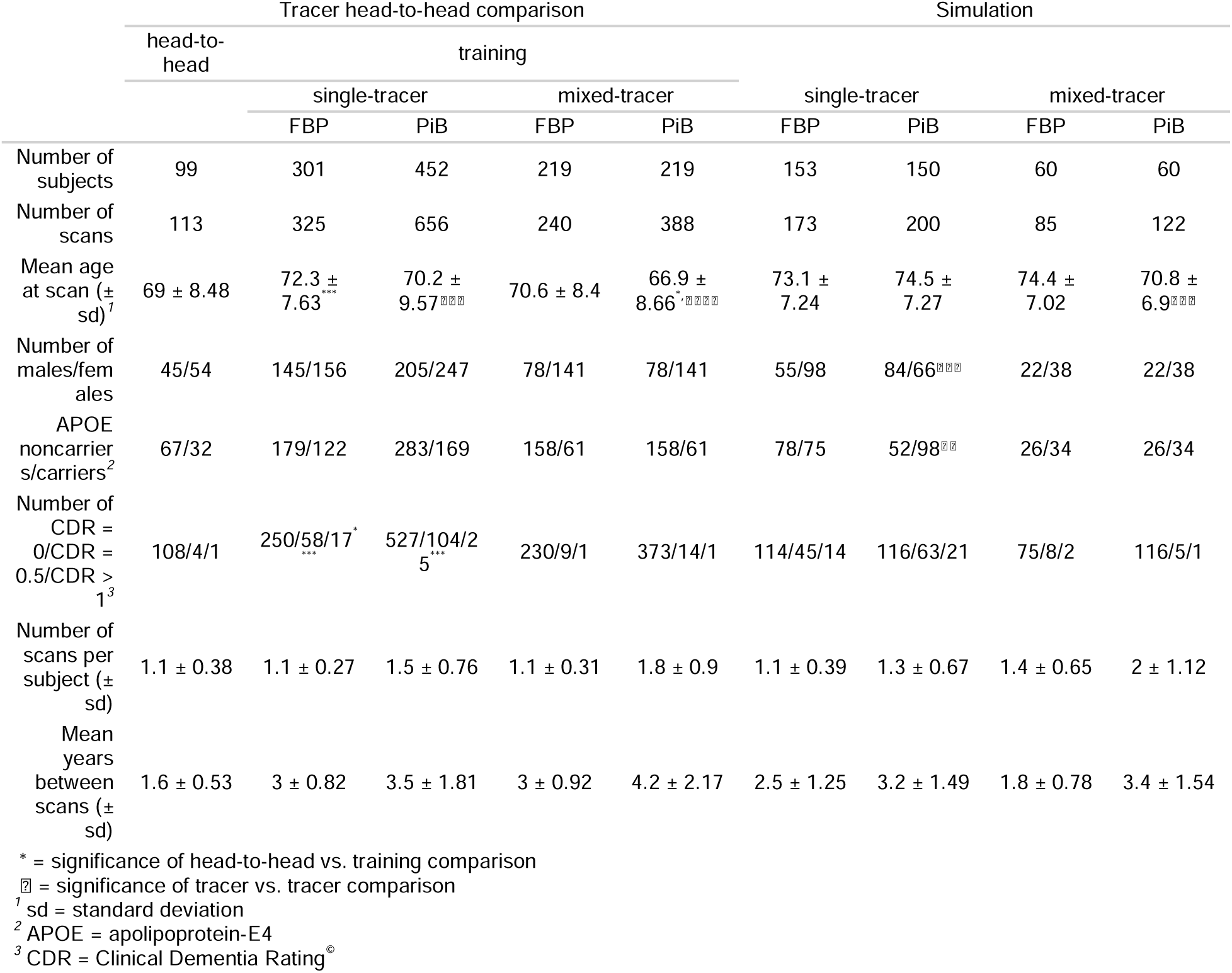
Demographics of each cohort. The training dataset of the head-to-head comparison was split into subjects who were scanned with only one tracer (single-tracer) and those who were scanned with multiple tracers (mixed-tracer). Statistically significant differences are denoted with asterisks and crosses. Asterisks indicate comparisons between head-to-head and each of the 4 training sub-cohorts, whereas crosses indicate FBP vs. PiB comparisons within a single/mixed tracer cohort. The number of symbols indicates the significance level (1 = p < 0.05, 2 = p < 0.01, 3 = p < 0.005, 4 = p < 1e-4). Note that the reported CDRs are counted by scans, where the closest CDR score in time was assigned to every scan.

### Tracer head-to-head comparison

We evaluated the ability of Centiloid, ComBat and PEACE to improve the absolute agreement between FBP and PiB using the head-to-head dataset. For the global summary region, absolute agreement, as measured by ICC, increased after harmonization with either Centiloid (/CC = 0.912) or ComBat with no covariates (/CC = 0.916), compared to the unharmonized SUVR (/CC = 0.882) (Table 2, Fig. 2). ICC also increased slightly after harmonization with ComBat with covariates () and GAM-ComBat (), albeit not to the same degree as ComBat with no covariates. Similarly, PEACE, either with (/CC = 0.884) or without (/CC = 0.897) covariates, did not perform as well as ComBat with no covariates or Centiloid in increasing ICC (Table 2, Supp. Fig. S1). For ROI measurements, all three ComBat harmonization methods led to a statistically significant increase in average ICC among all ROIs compared to unharmonized SUVR (p < 1e-4), with ComBat with no covariates again performing the best 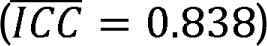 (Table 2, Fig. 3a). Additionally, ComBat with no covariates performed the best within the summary cortical ROIs 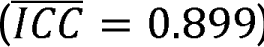 and other cortical ROIs 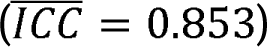 (Table 2, Fig. 3b-c). No method was effective at improving across-tracer agreement for the subcortical ROIs, with mean ICC of less than 0.75 even after harmonization (Table 2, Fig. 3d). PEACE did not improve the mean ICC with statistical significance in any of the groups except for other cortical ROIs (Supp. Fig. S2), and PEACE both with and without covariates achieved a lower mean ICC than ComBat with no covariates (Table 2). When plotting region-wise ICC on the inflated brain surface, we observed that Centiloid resulted in a decrease in ICC in the bilateral occipital and sensorimotor regions, and in the left temporal and parietal cortices (Supp. Fig. S3b). PEACE also exhibited a decrease in ICC in similar regions (Supp. Fig. S3f-g). In contrast, none of the ComBat variants led to such decrease in these regions (Supp. Fig. S3c-e), with ComBat with no covariates having the highest magnitude of change in ICC across multiple regions.

**Fig. 2.** Global summary measures computed from PiB and FBP scans in the tracer head-to-head dataset. The red line indicates the best fit line from ordinary least squares linear regression, the gray area indicates the confidence interval of the slope, and the black line represents the identity line. Intraclass correlation coefficient (ICC) is reported on the bottom right of each scatterplot.

**Table 2.** Mean ± standard deviation of ICC and MAE from the tracer head-to-head comparison. The mean across scan pairs was computed for the global summary region, whereas the mean across ROIs was computed for the FreeSurfer ROI. Bold values indicate the best performing harmonization method. Asterisks indicate statistical significance of paired t-tests comparing each harmonization method with unharmonized (* = p < 0.05, ** = p < 0.01, *** = p < 0.005, **** = p < 1e-4).

| Harmonization method | Global summary |  | FreeSurfer ROI |  |  |  |  |  |  |  |
| --- | --- | --- | --- | --- | --- | --- | --- | --- | --- | --- |
|  | ICC | MAE | All ROI |  | Summary cortical ROI |  | Other cortical ROI |  | Subcortical ROI |  |
|  |  |  | ICC | MAE | ICC | MAE | ICC | MAE | ICC | MAE |
| unharmonized | 0.882 | 0.101 ± 0.076 | 0.819 ± 0.077 | 0.111 ± 0.029 | 0.866 ± 0.03 | 0.113 ± 0.023 | 0.838 ± 0.048 | 0.109 ± 0.031 | 0.701 ± 0.089 | 0.12 ± 0.028 |
| Centiloid, scaled | 0.912 | 0.088 ± 0.075 | 0.811 ± 0.098 | 0.112 ± 0.032 | 0.891 ± 0.018 | 0.099 ± 0.02** | 0.822 ± 0.063 | 0.116 ± 0.037 | 0.686 ± 0.137 | 0.11 ± 0.019 |
| ComBat, no covariates | **0.916** | **0.08 ± 0.075** | **0.838 ± 0.083** **** | **0.086 ± 0.019** **** | **0.899 ± 0.018** ** | **0.089 ± 0.014** ** | **0.853 ± 0.05** **** | 0.084 ± 0.019**** | **0.72 ± 0.111** | **0.09 ± 0.021** |
| ComBat + age, sex, APOE | 0.898 | 0.099 ± 0.075 | 0.827 ± 0.08**** | 0.095 ± 0.023**** | 0.881 ± 0.024** | 0.106 ± 0.017 | 0.844 ± 0.049*** | 0.093 ± 0.024** | 0.707 ± 0.099 | 0.092 ± 0.022 |
| GAM-ComBat + age, sex, APOE | 0.897 | 0.1 ± 0.075 | 0.827 ± 0.08**** | 0.095 ± 0.023**** | 0.881 ± 0.024** | 0.107 ± 0.017 | 0.844 ± 0.049*** | 0.093 ± 0.024** | 0.707 ± 0.099 | 0.092 ± 0.022 |
| PEACE, no covariates | 0.897 | 0.092 ± 0.077** | 0.813 ± 0.087 | 0.097 ± 0.022**** | 0.875 ± 0.027 | 0.103 ± 0.019 | 0.831 ± 0.058 | 0.093 ± 0.021** | 0.684 ± 0.094 | 0.104 ± 0.027 |
| PEACE + age, sex, APOE | 0.884 | 0.083 ± 0.095** | 0.822 ± 0.079 | **0.086 ± 0.021** **** | 0.867 ± 0.028 | 0.094 ± 0.018 | 0.842 ± 0.05 | **0.083 ± 0.021** **** | 0.698 ± 0.087 | 0.092 ± 0.022 |

**Fig. 3.**
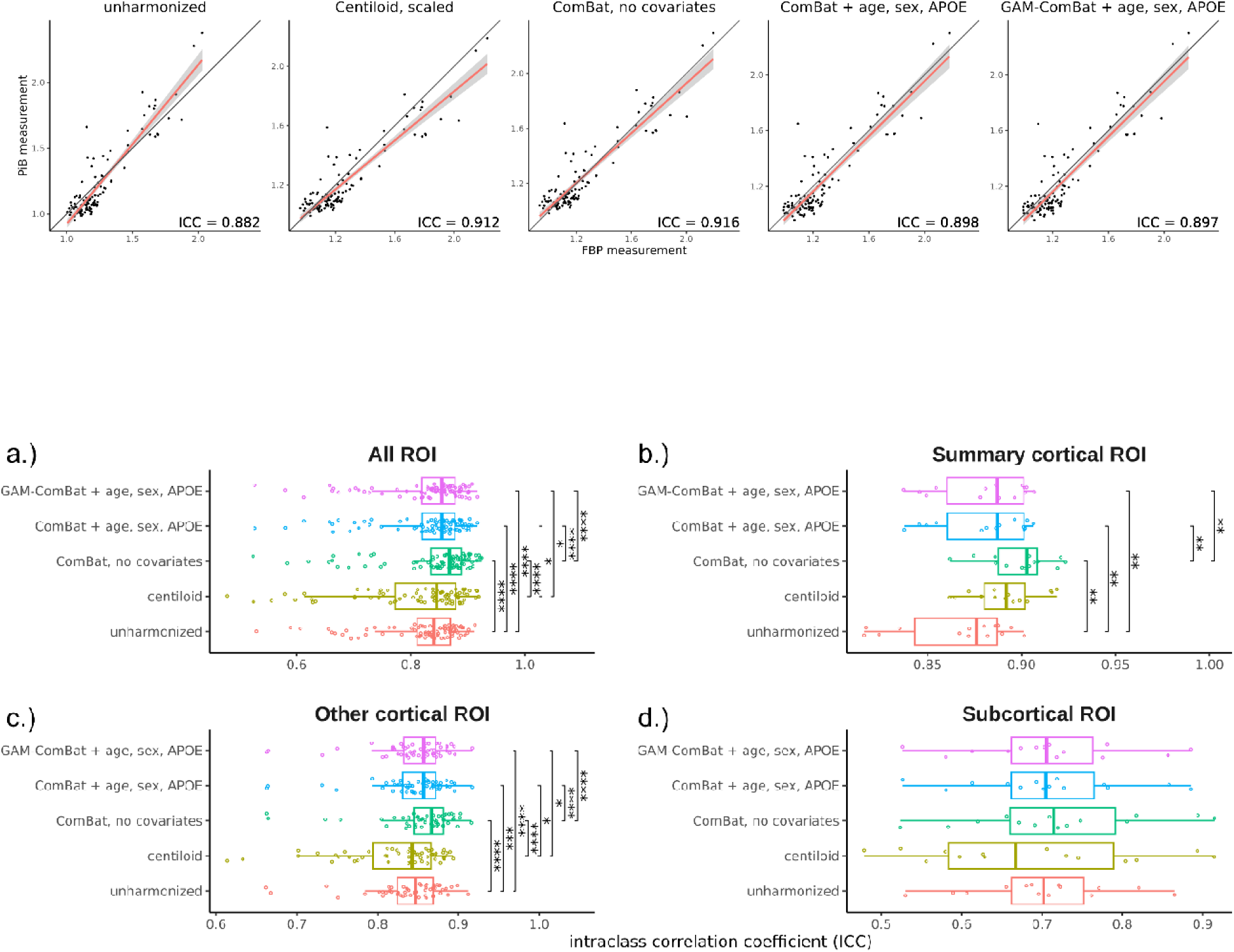
Distribution of regional ICC across all ROIs, grouped by harmonization method and ROI subgroup. Each point represents a single ROI. Significance levels from paired t-tests with Bonferroni correction are indicated for each pair of harmonization methods (* = p < 0.05, ** = p < 0.01, *** = p < 0.005, **** = p < 1e-4).

When assessing the absolute error between FBP and PiB measurements of the global summary region (Table 2, Fig. 4, Supp. Fig. S4), ComBat with no covariates (*MAE* = 0.080) and PEACE, both with (*MAE* = 0.083) and without (*MAE* = 0.092) covariates, were the only methods that reduced the mean absolute error with statistical significance (p < 0.05) compared to unharmonized SUVRs (*MAE* = 0.101). ComBat with no covariates lowered the absolute error the greatest between the three methods. For ROI measurements (Table 2, Fig. 5, Supp. Fig. S5), all methods except Centiloid significantly reduced the average MAE among all ROIs (p < 1e-4), but ComBat with no covariates and PEACE with covariates resulted in the greatest reduction 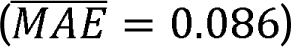. ComBat with no covariates also performed the best within the summary cortical ROIs 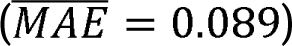 and subcortical ROIs 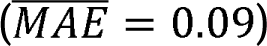, while PEACE with covariates performed the best within other cortical ROIs 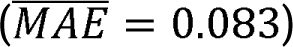, although ComBat with no covariates performed comparably. When plotting region-wise MAE on the inflated brain surface, we observed that both ComBat with no covariates and PEACE with covariates led to the greatest reduction in MAEs in ROIs across the bilateral frontal, parietal and occipital regions (Supp. Fig. S6).

**Fig. 4.**
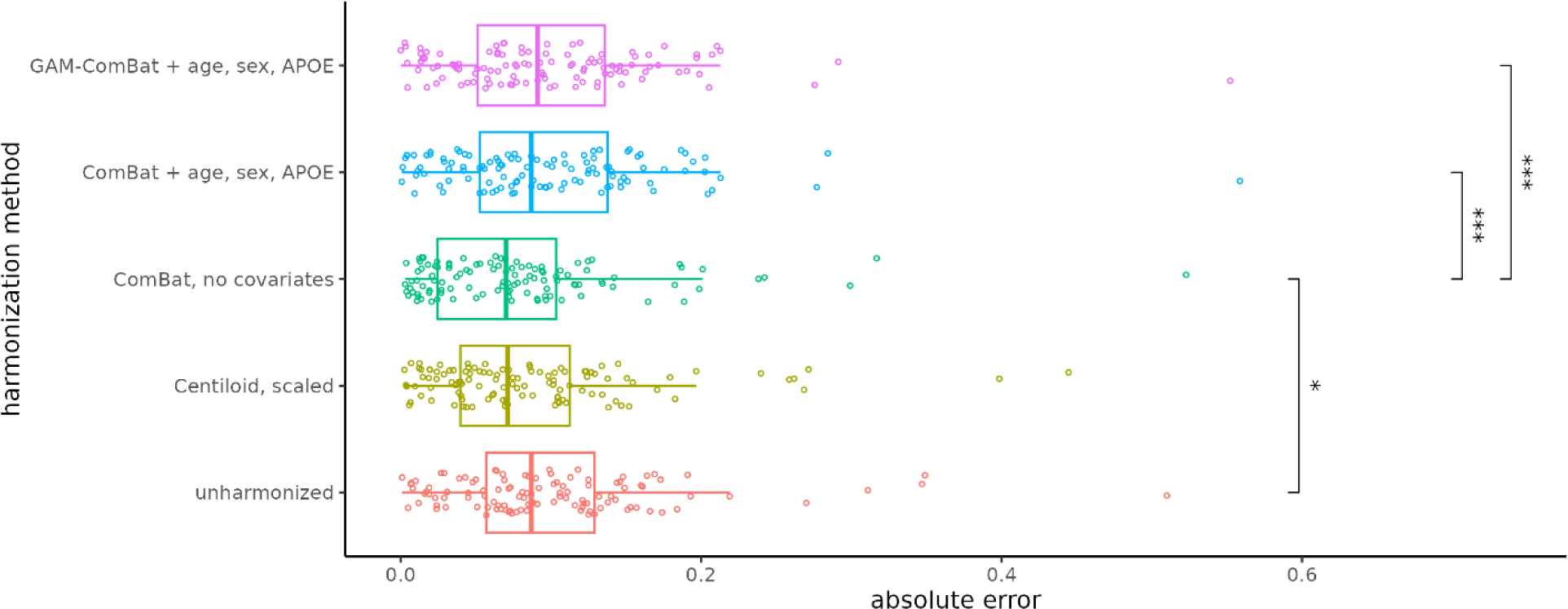
Absolute error of the global summary measure from each PiB and FBP scan pair in the tracer head-to-head dataset. Each point represents a single scan pair. Significance levels from paired t-tests with Bonferroni correction are indicated for each pair of harmonization methods (* = p < 0.05, ** = p < 0.01, *** = p < 0.005, **** = p < 1e-4).

**Fig. 5.**
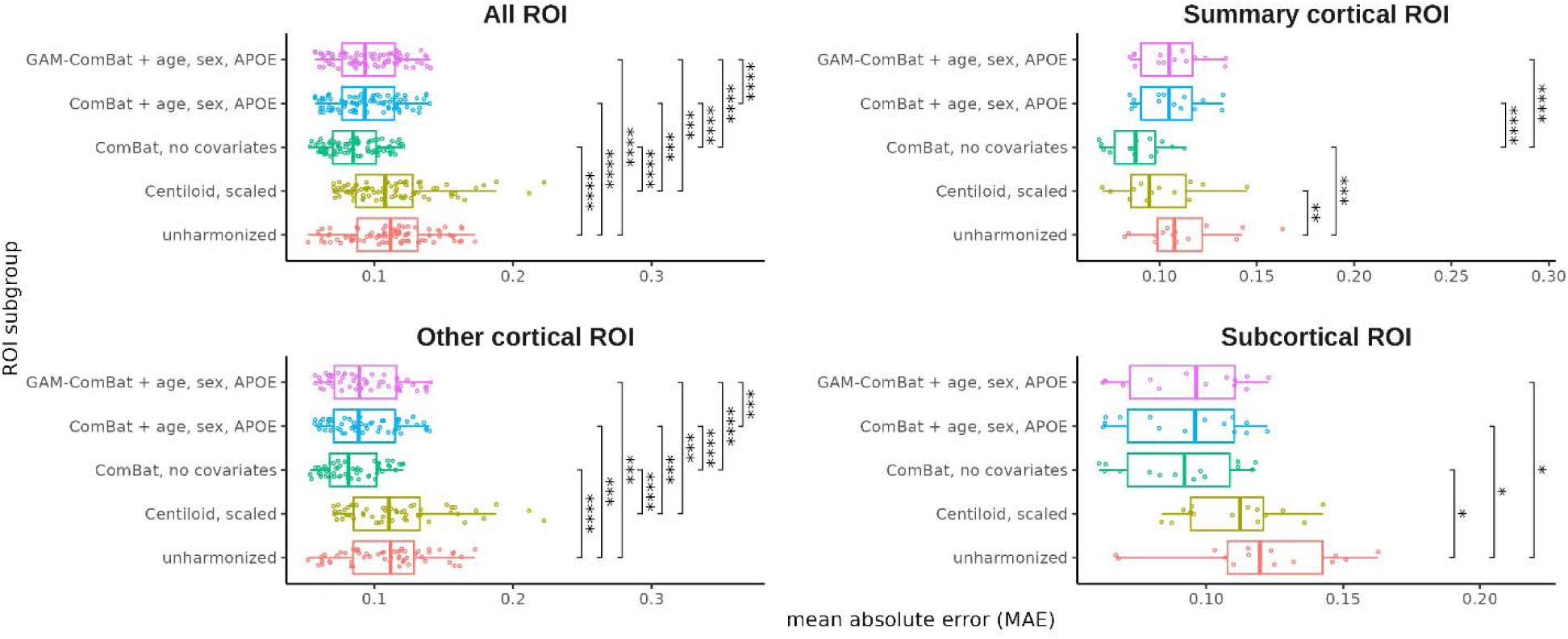
Distribution of regional MAE across all ROIs, grouped by harmonization method and ROI subgroup. Each point represents a single ROI. Significance levels from paired t-tests with Bonferroni correction are indicated for each pair of harmonization methods (* = p < 0.05, ** = p < 0.01, *** = p < 0.005, **** = p < 1e-4).

## Clinical trial simulation

We performed simulations to test for group differences in amyloid rate-of-change between treatment and placebo groups in a hypothetical clinical trial, and evaluated whether harmonization improved the ability of detecting these differences. For the global summary SUVR, both Centiloid and ComBat without covariates resulted in overall increases in statistical power after harmonization in the presence of a treatment effect (i.e., for rate-of-change E {-0.01, -0.02, -0.03}), primarily when the placebo group had high FBP composition and the treatment group had low FBP composition (Fig. 6). A similar increase in power was observed when using ComBat with covariates, albeit at a lesser magnitude. However, PEACE led to widespread decreases in power, and although longitudinal ComBat led to increases in power in certain tracer composition configurations, the effect was not as consistent as either Centiloid or ComBat with no covariates (Supp. Fig. S7). In the absence of a treatment effect (i.e., rate-of-change = 0), Centiloid, ComBat and longitudinal ComBat with no covariates achieved large decreases in Type-I error, primarily when the placebo group had low FBP composition and the treatment group had high FBP composition. Slight decreases in power were also observed in the case of a low treatment effect (i.e., rate-of-change = -0.01). Supplementary Table S2 shows the mean power computed across all 25 configurations of treatment and placebo FBP compositions. Out of all methods, Centiloid achieved the largest mean power for detecting the treatment effect in every rate-of-change except for -0.01 (longitudinal ComBat performed the best in this case), and Centiloid also achieved the lowest mean Type-I error. However, ComBat with no covariates was often the second-best performing method.

**Fig. 6.**
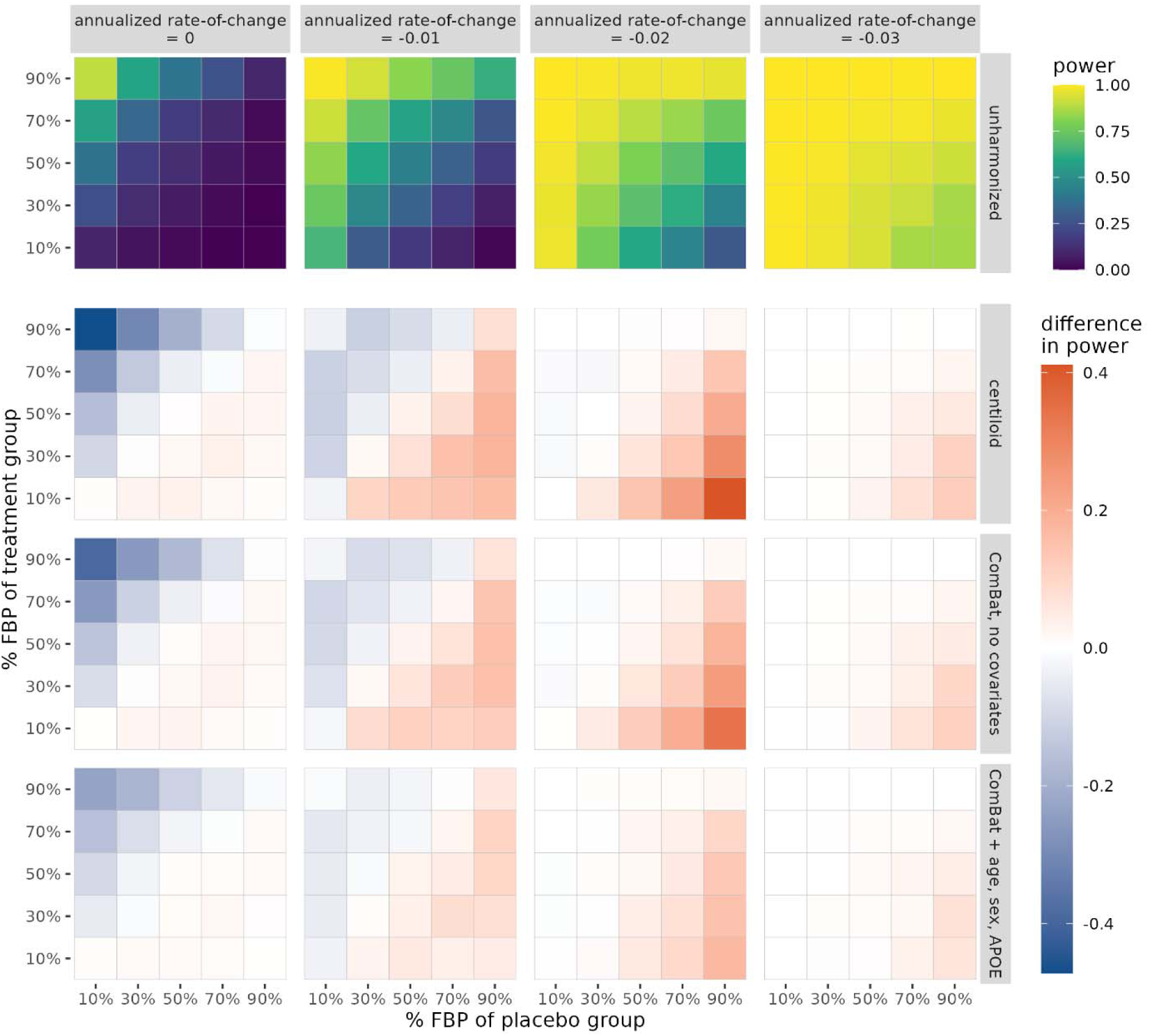
Statistical power of detecting group differences in rate-of-change of the global summary SUVR between treatment and placebo groups, computed as the proportion of significant findings over 1000 iterations. Power is plotted for unharmonized SUVR, while the difference in power relative to unharmonized is plotted for all harmonization methods. The true underlying rate-of-change is varied across columns. The proportion of FBP scans in the placebo and treatment groups is varied across the horizontal and vertical axes of each heatmap, respectively. Note that for annualized rate-of-change equal to zero, the proportion of significant findings corresponds to Type-I error rate.

For regional SUVRs, similar patterns of change in power after harmonization were observed. Across all ROIs, mean power was increased/Type-I error was decreased after harmonization with either Centiloid or ComBat, with both methods producing comparable changes in power (Supp. Table S3). Additionally, these same changes in regional power after harmonization were consistent across all three ROI subgroups. Surface plots revealed that regions which exhibited relatively low power of detecting treatment effects (such as the frontal and medial parietal cortices and putamen in the case of rate-of-change = -0.02) experienced a high increase in power after harmonization with either Centiloid or ComBat with no covariates (Supp. Fig. S8-9). Again, harmonization with PEACE led to widespread decreases in statistical power across multiple regions, and longitudinal ComBat showed worse performance in increasing power compared to Centiloid or ComBat with no covariates, with the exception of when rate-of-change = -0.01 (Supp. Fig. S8-9, Supp. Table S3).

## Discussion

We demonstrated that ComBat may effectively harmonize amyloid PET measurements across FBP and PiB. Notably, ComBat with no covariates outperformed Centiloid in increasing absolute agreement between tracers in both the global summary and regional measurements, and resulted in a comparable improvement in detecting group differences in the simulated clinical trial. As more studies shift focus from using a global summary metric of amyloid burden to using the spatial distribution of regional amyloid as features [6,7], harmonization techniques like ComBat that can be applied to multiple regions become appealing for pooling PET data across multiple tracers.

ComBat poses several methodological advantages over Centiloid. Firstly, calibration of Centiloid requires *a priori* selection of representative individuals from dichotomous groups, namely healthy control and typical AD cohorts. This *a priori* cohort selection may introduce bias into the calibration process. Especially if the selected sample is small and/or captures only a subset of the overall population (e.g., biased towards a single ethnicity group), Centiloid may not generalize well to heterogeneous or out-of-sample datasets. ComBat circumvents this requirement, which allows it to learn a robust harmonization on a wider spectrum of data which consists of controls, AD patients and “in-between” subjects. Furthermore, much like Centiloid, a trained ComBat model may then be used to harmonize out-of-sample data. However, one should still carefully consider the subject selection process and ensure that the proportions of cohorts are not skewed towards a particular cohort (e.g., many more healthy controls than AD patients), since this may lead to a suboptimal harmonization model [29]. Secondly, Centiloid requires at least two PET scans of different tracers for each subject in the calibration cohort, one of which should be acquired using PiB. In contrast, ComBat can train using just one scan per subject and does not require PiB to be used. Indeed, we expect ComBat to generalize well to harmonizing two (or more) [^18^F]-based tracers without the need for PiB, although further investigation using head-to-head data of these tracers should be conducted to verify this. Thirdly, a region-specific harmonization is important for addressing sources of tracer bias which variably affect different regions, such as non-specific binding [35]. However, as suggested by Klunk et al. [18], a region-specific Centiloid calibration is not ideal, since it would fix different SUVRs of different regions to the same Centiloid value. In contrast, ComBat independently removes the tracer-specific variance from the target measurements without scaling the dynamic range of each region to fixed points, making it a more suitable technique for regional harmonization. A caveat to this is that ComBat effectively centers the harmonized measurements on the global mean and variance of the data. As such, these measurements can no longer be interpreted as belonging to the scale of a particular batch, but rather on an aggregated scale. Alternative approaches such as modified ComBat [36] allow users to choose a “gold standard” reference batch to which all other batches are adjusted to, which may aid in improving the interpretability of harmonized measurements.

Lastly, ComBat has the advantage of being able to preserve covariate relationships in the target measurement, which may be useful in downstream analyses such as in predictive models which take into account biologically-related variance to make accurate predictions. However, in the context of purely evaluating the absolute agreement between tracers, we observed that including covariates into the ComBat model led to worse ICC and absolute errors. This may be partially due to differences in covariate distributions between the training and head-to-head cohorts, of which age and CDR differed with statistical significance. Although CDR was not explicitly included as a ComBat covariate, it may have indirectly contributed to a biased ComBat model which does not generalize well to testing data with different covariate characteristics.

It was noted that no harmonization method investigated in this study performed well for the subcortical regions. Notably, these regions lie close to white matter regions, and thus may be affected by non-specific binding more so than cortical regions. This may contribute to more noise in the subcortical regions, which batch harmonization methods such as ComBat are not able to mitigate. One potential area of investigation is to evaluate whether partial volume correction [35] would have an effect on regional harmonization of PET SUVRs, especially for regions which experience high amounts of signal spill-over from neighboring white matter regions.

Our simulation experiments revealed the importance of harmonization in settings where multiple tracers are utilized to track brain amyloid deposition in clinical trial participants. Particularly, harmonization was the most beneficial when trial groups exhibited differing proportions of tracer data. In these “off-diagonal” cases, tracer biases contributed to a substantial confounding effect across clinical trial groups, resulting in either a reduction of power in detecting the true underlying treatment effect, or an increase in Type-I error in the case when no treatment effect exists. Harmonization effectively served to mitigate these confounding effects due to tracer differences. This was consistent with previous reports that found significant differences in amyloid rates-of-change across different tracers within real clinical trial groups, and that these differences were subsequently removed after harmonization [17]. Interestingly, we observed an asymmetric effect where harmonization led to changes in power in one off-diagonal, but not in the other. This was most likely because we utilized a one-sided statistical test to test for rate-of-change differences. In the case of low FBP% in the placebo group and high FBP% in the treatment group, and when there was no ground-truth treatment effect introduced, a high amount of Type-I error suggested that FBP contributed to a greater rate-of-change compared to PiB due to tracer biases alone. However, in the opposite off-diagonal, these biases did not contribute to any Type-I error. While tracer biases were still present in the overall data, this indicated that they did not interfere with the detection of one-way group differences.

It is important to note, however, that scenarios of high imbalance of tracer proportions between clinical trial arms are very unlikely to occur in a real-world setting, assuming proper randomization. In the more realistic case where trial groups exhibit an equal proportion of tracer data, a much lower change in power was observed compared to the off-diagonal cases. This is likely because the same tracer bias would affect both groups equally, which statistically would not influence the detection of group differences.

We investigated PEACE and longitudinal ComBat, which were previously validated for scanner-wise harmonization, for specifically tracer harmonization in the current study. Both methods led to mixed results in the head-to-head comparison as well as in the clinical trial simulation. In the head-to-head comparison, PEACE with covariates performed similarly to ComBat with no covariates in terms of MAE, but not in terms of ICC. Additionally, in the clinical trial simulation, PEACE failed to improve statistical power of detecting the treatment effect. This can partly be explained by our decision to simulate SUVRs following a unimodal Gaussian distribution, which was motivated by the fact that a true clinical trial will only enroll amyloid-positive participants and exclude amyloid-negative individuals. Longitudinal ComBat resulted in mixed improvements in power, but these improvements were not as consistent compared to Centiloid or ComBat with no covariates. We speculate that increased model complexity may have contributed to models which were less robust to the data at hand. Ultimately, we found that ComBat with no covariates, which is the simplest model with the fewest number of parameters to estimate and the fewest assumptions made, consistently performed either comparably to or better than PEACE or longitudinal ComBat.

There are several limitations to the current work. Firstly, on the basis of purely increasing tracer agreement, there are no clear recommendations on the choice of including covariates in ComBat. One caveat to using ComBat is that, unless explicitly accounted for in the covariates, it will assume that any biases due to real biological differences between tracer cohorts are batch differences, which are subsequently removed. Therefore, one should carefully examine the composition of the data at hand and consider whether it is necessary to model known biological factors via the covariate terms. Secondly, data from the simulation experiment were generated from models trained on a cohort of amyloid-positive subjects from OASIS-3 instead of data from an actual anti-amyloid drug trial. Although simulations were set up to mimic data that would be collected in a successful trial, it remains to be seen whether our hypotheses would hold on real-world clinical trial data. Thirdly, our conclusions on the performance of ComBat for tracer harmonization are limited to PiB and FBP. Although we expect ComBat to be robust to other ^18^F-based amyloid tracers such as [^18^F]-florbetaben and [^18^F]-flutemetamol, future work is required to validate this using head-to-head data from these tracers. Finally, our simulation analysis only focused on early amyloid-positive individuals, which we assumed to exhibit temporal amyloid accumulation in a roughly linear fashion. However, to draw conclusions on a cohort of both amyloid-negative and positive individuals (and even late-stage individuals with plateaued amyloidosis), a sigmoidal or piecewise linear model should instead be used in order to model the non-linearities of amyloid accumulation across the broader AD spectrum [37,38].

## Conclusion

Harmonization of amyloid PET radiotracers is imperative for removing tracer-specific biases in amyloid burden measurements for optimal performance of downstream tasks, such as enhancing statistical power and reducing false discoveries in clinical trials. In the current study, we demonstrated that ComBat is effective for harmonizing both global and regional amyloid measurements in an entirely data-driven way. Our experimental results suggest that ComBat not only increases the absolute agreement of measurements made within scan pairs of the same group of subjects by different tracers, but also provides a significant benefit to the performance of detecting true treatment effects in anti-amyloid drug trials. ComBat thus presents as a viable technique for harmonizing regional-based analyses of amyloid PET.

## Supplementary Information

**Additional file 1.docx** - Supplementary figures and tables. **Supp. Table S1**: List of FreeSurfer regions-of-interest used in the study, along with their subgroupings. **Supp. Fig. S1**: Scatterplots of the global summary SUVR from the tracer head-to-head comparison, with results of PEACE. **Supp. Fig. S2**: Boxplots of regional SUVRs from the tracer head-to-head comparison, with results of PEACE. **Supp. Fig. S3**: Regional ICCs plotted on the surface from the tracer head-to-head comparison. **Supp. Fig. S4**: Boxplots of absolute errors of the global summary SUVR from the tracer head-to-head comparison, with results of PEACE. **Supp. Fig. S5**: Boxplots of mean absolute errors of the regional SUVR from the tracer head-to-head comparison, with results of PEACE. **Supp. Fig. S6**: Regional MAEs plotted on the surface from the tracer head-to-head comparison. **Supp. Fig. S7**: Heatmaps of statistical power of the clinical trial simulation when run on the global summary SUVR, with results of PEACE and longitudinal ComBat. **Supp. Fig. S8**: Brain surface plots of mean statistical power from the simulation experiment. **Supp. Fig. S9**: Subcortical plots of mean statistical power from the simulation experiment. **Supp. Table S2**: Mean statistical power of detecting significant rate-of-change differences between treatment and placebo groups in the simulation experiment for the global summary amyloid estimate. **Supp. Table S3**: Mean statistical power of detecting significant rate-of-change differences between treatment and placebo groups in the simulation experiment for the ROI amyloid measurements.

## Key Points

- ComBat is a data driven harmonization method which, unlike Centiloid, does not require *a priori* selection and stratification of training cohorts and is able to harmonize regional amyloid PET estimates.
- ComBat with no covariates performed the best in increasing the absolute agreement of regional amyloid PET measurements made within scan pairs of the same group of subjects using two different radiotracers.
- ComBat increased the statistical power of detecting treatment effects and decreased Type-I error of falsely detecting effects in a simulated anti-amyloid drug trial.

## Declarations

### Ethics approval and consent to participate

Ethics approvals were obtained by the OASIS-3 dataset. All participants were consented into Knight ADRC-related projects in accordance with the Declaration of Helsinki and following procedures approved by the Institutional Review Board of Washington University School of Medicine in St. Louis. For more details, we refer the reader to the OASIS-3 reference.

### Consent for publication

Not applicable

### Availability of data and materials

Data utilized in this study were obtained from the OASIS-3 open access dataset. Data can be requested at https://sites.wustl.edu/oasisbrains.

Code for this study will be made publicly available at https://github.com/sotiraslab. All statistical analyses and simulation experiments were implemented using R version 4.4.0 and python version 3.10.10. The *neuroharmonize* python package (https://github.com/rpomponio/neuroHarmonize) was used to train and apply ComBat and GAM-ComBat models, and the *longCombat* R package (https://github.com/jcbeer/longCombat) was used to train and apply longitudinal ComBat.

### Authors’ contributions

All authors contributed to the conceptualization and design of the study. BY implemented all data analyses and experiments and wrote the first draft of the manuscript. AS, BG and TB contributed to the interpretation of data. TE, SK and DK provided technical support. All authors were involved with manuscript revision, and all approved of the final draft.

## Data Availability

Data utilized in this study were obtained from the OASIS-3 open access dataset. Data can be requested at https://sites.wustl.edu/oasisbrains.
Code for this study will be made publicly available at https://github.com/sotiraslab. All statistical analyses and simulation experiments were implemented using R version 4.4.0 and python version 3.10.10. The neuroharmonize python package (https://github.com/rpomponio/neuroHarmonize) was used to train and apply ComBat and GAM-ComBat models, and the longCombat R package (https://github.com/jcbeer/longCombat) was used to train and apply longitudinal ComBat.

## Acknowledgements

Acknowledgement is made to the donors of the ADR A2021042S, a program of the BrightFocus Foundation, for support of this research. Data were provided by OASIS-3: Longitudinal Multimodal Neuroimaging (Principal Investigators: T. Benzinger, D. Marcus, J. Morris). OASIS-3 was supported by the following funding sources: NIH P50 AG00561, P30 NS09857781, P01 AG026276, P01 AG003991, R01 AG043434, UL1 TR000448, R01 EB009352. AV-45 doses were provided by Avid Radiopharmaceuticals, a wholly owned subsidiary of Eli Lilly.

## List of Abbreviations

PET: positron emission tomography
FBP: [^18^F]-florbetapir
PiB: [^11^C]-Pittsburgh Compound-B
OASIS-3: Open Access Series of Imaging Studies 3
SUVR: standardized uptake value ratio
ICC: intraclass correlation coefficient
MAE: mean absolute error
AD: Alzheimer’s disease
ROI: region-of-interest
CL: Centiloid
MRI: magnetic resonance imaging
APOE: apolipoprotein-ε4
GAM: generalized additive model
LME: linear mixed effects
CDR: Clinical Dementia Rating
PEACE: Probabilistic Estimation for Across-batch Compatibility Enhancement.

