## Supplementary Material for "Evaluation of ComBat harmonization for reducing across-tracer differences in regional amyloid PET analyses"

**Supplementary Table S1** List of FreeSurfer regions-of-interest (ROI) used in the study, along with their subgroupings.

| **FreeSurfer ROI** | **ROI subgroup** |
| --- | --- |
| ctx.lh.bankssts | Other cortical ROI |
| ctx.lh.caudalanteriorcingulate | Other cortical ROI |
| ctx.lh.caudalmiddlefrontal | Other cortical ROI |
| ctx.lh.cuneus | Other cortical ROI |
| ctx.lh.entorhinal | Other cortical ROI |
| ctx.lh.frontalpole | Other cortical ROI |
| ctx.lh.fusiform | Other cortical ROI |
| ctx.lh.inferiorparietal | Other cortical ROI |
| ctx.lh.inferiortemporal | Other cortical ROI |
| ctx.lh.insula | Other cortical ROI |
| ctx.lh.isthmuscingulate | Other cortical ROI |
| ctx.lh.lateraloccipital | Other cortical ROI |
| ctx.lh.lingual | Other cortical ROI |
| ctx.lh.paracentral | Other cortical ROI |
| ctx.lh.parahippocampal | Other cortical ROI |
| ctx.lh.parsopercularis | Other cortical ROI |
| ctx.lh.parsorbitalis | Other cortical ROI |
| ctx.lh.parstriangularis | Other cortical ROI |
| ctx.lh.pericalcarine | Other cortical ROI |
| ctx.lh.postcentral | Other cortical ROI |
| ctx.lh.posteriorcingulate | Other cortical ROI |
| ctx.lh.precentral | Other cortical ROI |
| ctx.lh.rostralanteriorcingulate | Other cortical ROI |
| ctx.lh.superiorparietal | Other cortical ROI |
| ctx.lh.supramarginal | Other cortical ROI |
| ctx.lh.temporalpole | Other cortical ROI |
| ctx.lh.transversetemporal | Other cortical ROI |
| ctx.rh.bankssts | Other cortical ROI |
| ctx.rh.caudalanteriorcingulate | Other cortical ROI |
| ctx.rh.caudalmiddlefrontal | Other cortical ROI |
| ctx.rh.cuneus | Other cortical ROI |
| ctx.rh.entorhinal | Other cortical ROI |
| ctx.rh.frontalpole | Other cortical ROI |
| ctx.rh.fusiform | Other cortical ROI |
| ctx.rh.inferiorparietal | Other cortical ROI |
| ctx.rh.inferiortemporal | Other cortical ROI |
| ctx.rh.insula | Other cortical ROI |
| ctx.rh.isthmuscingulate | Other cortical ROI |
| ctx.rh.lateraloccipital | Other cortical ROI |
| ctx.rh.lingual | Other cortical ROI |
| ctx.rh.paracentral | Other cortical ROI |
| ctx.rh.parahippocampal | Other cortical ROI |
| ctx.rh.parsopercularis | Other cortical ROI |
| ctx.rh.parsorbitalis | Other cortical ROI |
| ctx.rh.parstriangularis | Other cortical ROI |
| ctx.rh.pericalcarine | Other cortical ROI |
| ctx.rh.postcentral | Other cortical ROI |
| ctx.rh.posteriorcingulate | Other cortical ROI |
| ctx.rh.precentral | Other cortical ROI |
| ctx.rh.rostralanteriorcingulate | Other cortical ROI |
| ctx.rh.superiorparietal | Other cortical ROI |
| ctx.rh.supramarginal | Other cortical ROI |
| ctx.rh.temporalpole | Other cortical ROI |
| ctx.rh.transversetemporal | Other cortical ROI |
| left.cerebellum.cortex | Reference ROI |
| right.cerebellum.cortex | Reference ROI |
| left.accumbens.area | Subcortical ROI |
| left.amygdala | Subcortical ROI |
| left.caudate | Subcortical ROI |
| left.hippocampus | Subcortical ROI |
| left.pallidum | Subcortical ROI |
| left.putamen | Subcortical ROI |
| left.thalamus | Subcortical ROI |
| right.accumbens.area | Subcortical ROI |
| right.amygdala | Subcortical ROI |
| right.caudate | Subcortical ROI |
| right.hippocampus | Subcortical ROI |
| right.pallidum | Subcortical ROI |
| right.putamen | Subcortical ROI |
| right.thalamus | Subcortical ROI |
| ctx.lh.lateralorbitofrontal | Summary cortical ROI |
| ctx.lh.medialorbitofrontal | Summary cortical ROI |
| ctx.lh.middletemporal | Summary cortical ROI |
| ctx.lh.precuneus | Summary cortical ROI |
| ctx.lh.rostralmiddlefrontal | Summary cortical ROI |
| ctx.lh.superiorfrontal | Summary cortical ROI |
| ctx.lh.superiortemporal | Summary cortical ROI |
| ctx.rh.lateralorbitofrontal | Summary cortical ROI |
| ctx.rh.medialorbitofrontal | Summary cortical ROI |
| ctx.rh.middletemporal | Summary cortical ROI |
| ctx.rh.precuneus | Summary cortical ROI |
| ctx.rh.rostralmiddlefrontal | Summary cortical ROI |
| ctx.rh.superiorfrontal | Summary cortical ROI |
| ctx.rh.superiortemporal | Summary cortical ROI |

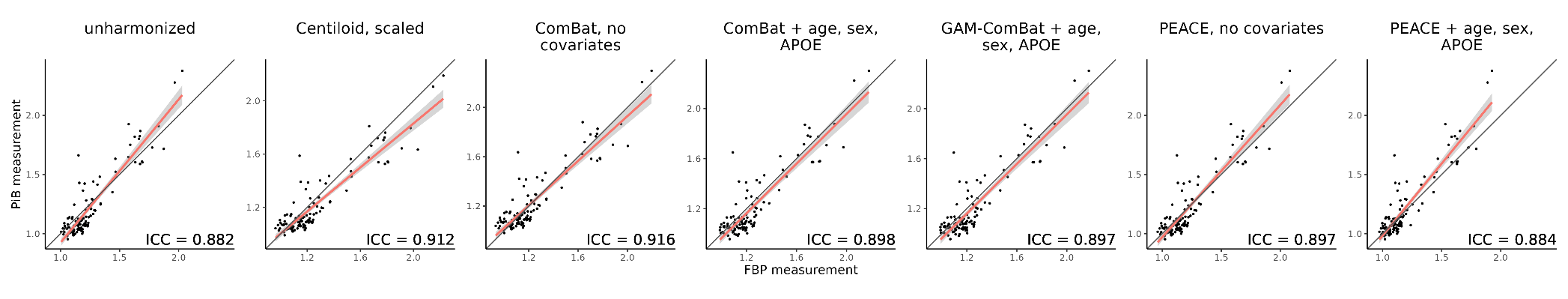

**Supplementary Fig. S1** Global summary measures computed from PiB and FBP scans in the tracer head-to-head dataset. The red line indicates the best fit line from ordinary least squares linear regression, the gray area indicates the confidence interval of the slope, and the black line represents the identity line. Intraclass correlation coefficient (ICC) is reported on the bottom right of each scatterplot. Plots for unharmonized SUVR, Centiloid, ComBat without covariates, ComBat with covariates, and GAM-ComBat are the same as Fig. 2 of the main text. Results for PEACE are also shown in the right two plots.

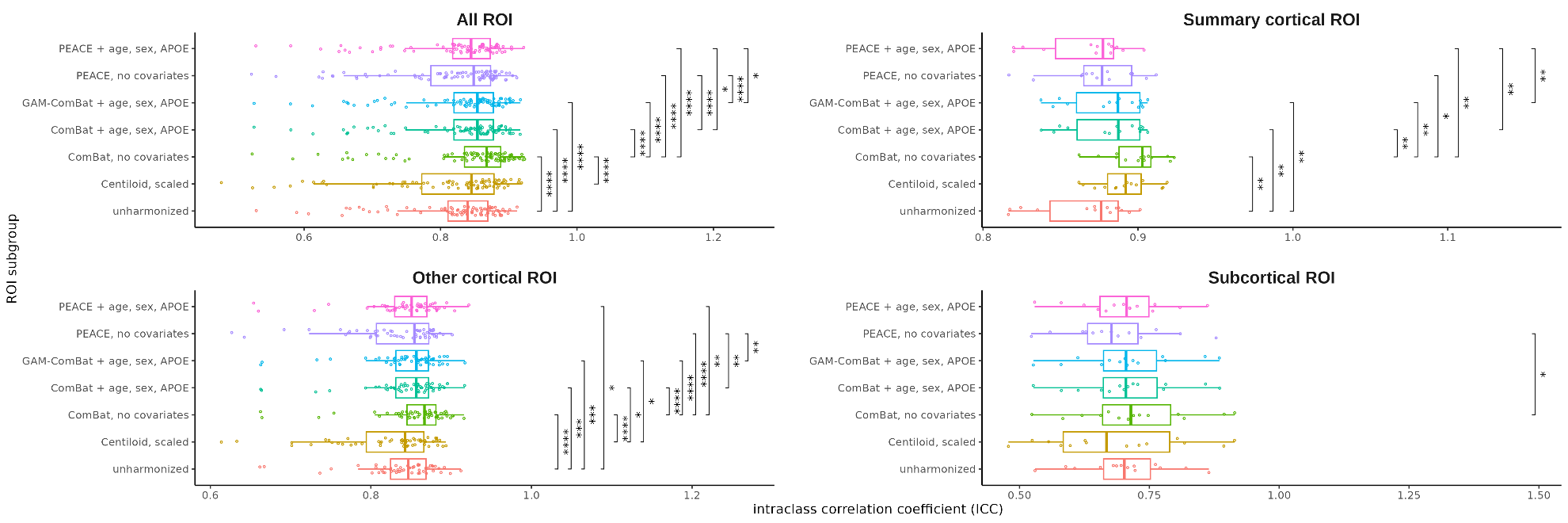

**Supplementary Fig. S2** Distribution of regional ICC across all ROIs, grouped by harmonization method and ROI subgroup. Each point corresponds to a single ROI. Significance levels from paired t-tests with Bonferroni correction are indicated above each pair of harmonization methods (* = p < 0.05, ** = p < 0.01, *** = p < 0.005, **** = p < 1e-4). Boxplots for unharmonized SUVR, Centiloid, ComBat without covariates, ComBat with covariates, and GAM-ComBat are the same as Fig. 3 of the main text. Results for PEACE are also shown in this figure.

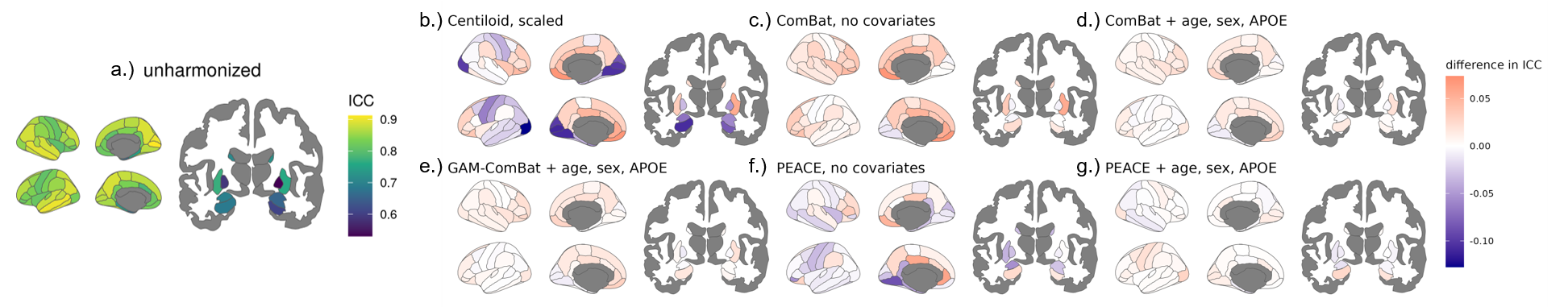

**Supplementary Fig. S3** Regional ICCs from the tracer head-to-head comparison experiments are plotted on the surface. (a) ICC values for unharmonized SUVR data are shown. The difference in ICC relative to the unharmonized data is plotted for each harmonization method: (b) Centiloid; (c) ComBat without covariates; (d) ComBat with age, sex and APOE-ε4 as linear covariates; (e) GAM-ComBat with age as a non-linear covariate and sex and APOE-ε4 as linear covariates; (f) PEACE without covariates; (g) PEACE with age, sex and APOE-ε4 as linear covariates.

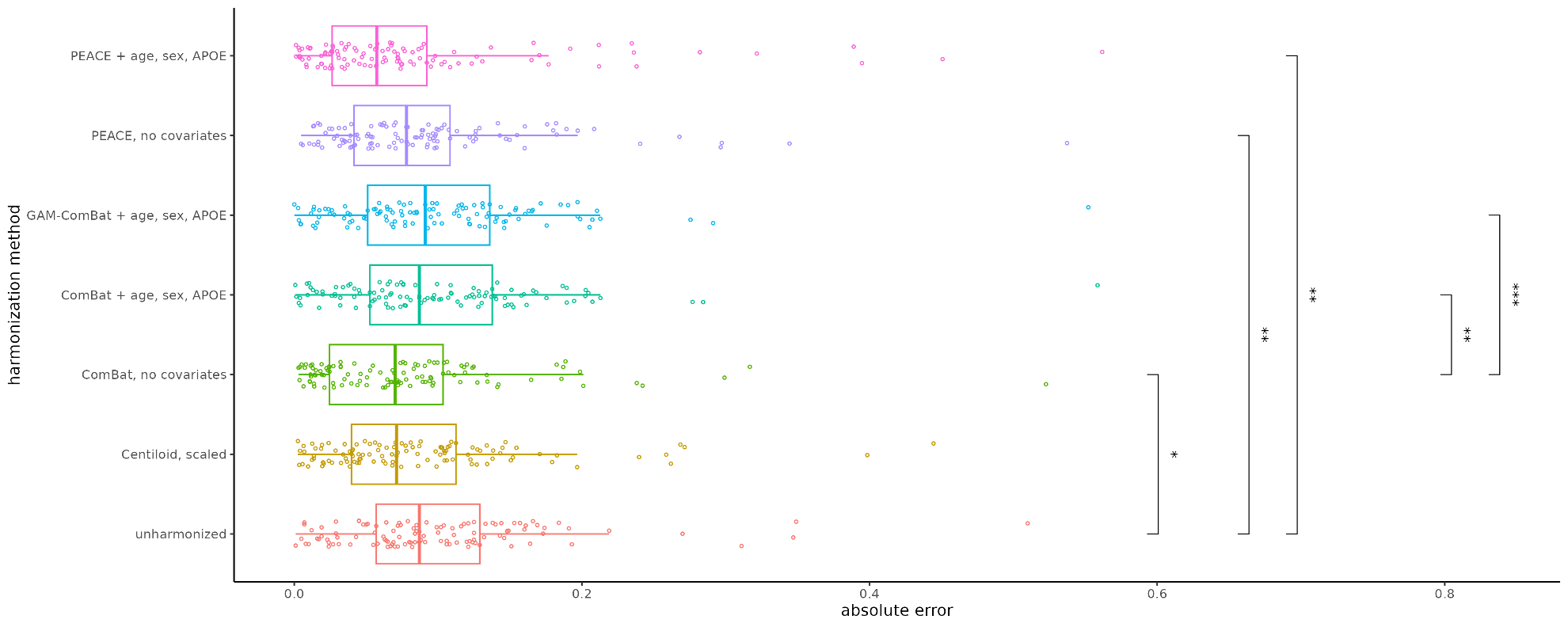

**Supplementary Fig. S4** Absolute error of the global summary measure from each PiB and FBP scan pair in the tracer head-to-head dataset. Each point represents a single scan pair. Significance levels from paired t-tests with Bonferroni correction are indicated for each pair of harmonization methods (* = p < 0.05, ** = p < 0.01, *** = p < 0.005, **** = p < 1e-4). Boxplots for unharmonized SUVR, Centiloid, ComBat without covariates, ComBat with covariates, and GAM-ComBat are the same as Fig. 4 of the main text. Results for PEACE are also shown in this figure.
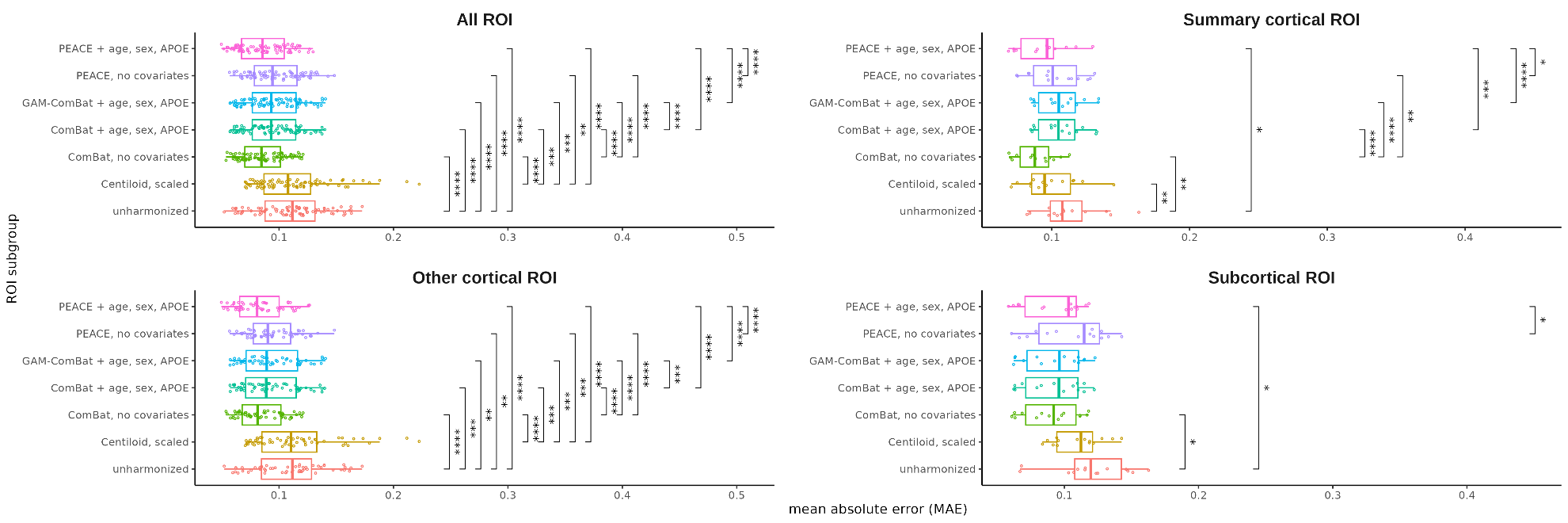

**Supplementary Fig. S5** Distribution of regional MAE across all ROIs, grouped by harmonization method and ROI subgroup. Each point represents a single ROI. Significance levels from paired t-tests with Bonferroni correction are indicated for each pair of harmonization methods (* = p < 0.05, ** = p < 0.01, *** = p < 0.005, **** = p < 1e-4). Boxplots for unharmonized SUVR, Centiloid, ComBat without covariates, ComBat with covariates, and GAM-ComBat are the same as Fig. 5 of the main text. Results for PEACE are also shown in this figure.

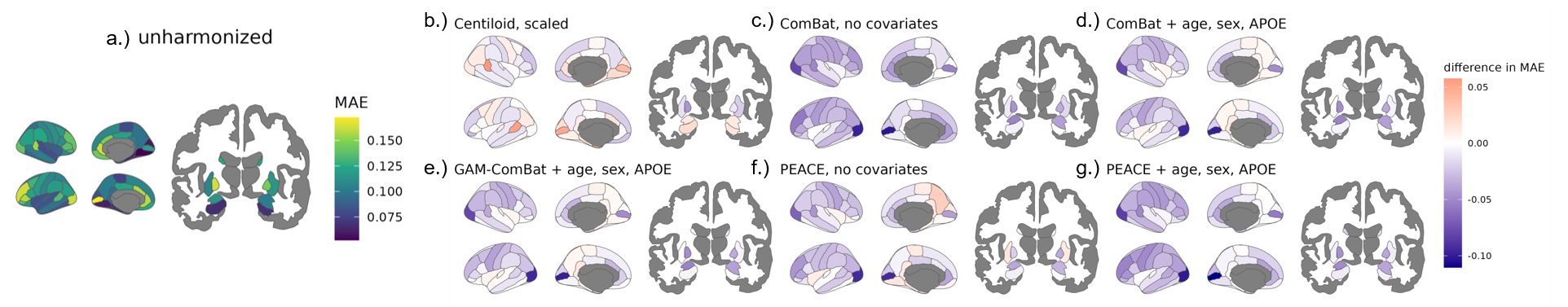

**Supplementary Fig. S6** Regional MAE values from the tracer head-to-head comparison experiment are plotted on the surface. MAE values for unharmonized SUVR data are shown. The difference in MAE relative to the unharmonized data is plotted for each harmonization method: (b) Centiloid; (c) ComBat without covariates; (d) ComBat with age, sex and APOE-ε4 as linear covariates; (e) GAM-ComBat with age as a non-linear covariate and sex and APOE-ε4 as linear covariates; (f) PEACE without covariates; (g) PEACE with age, sex and APOE-ε4 as linear covariates.

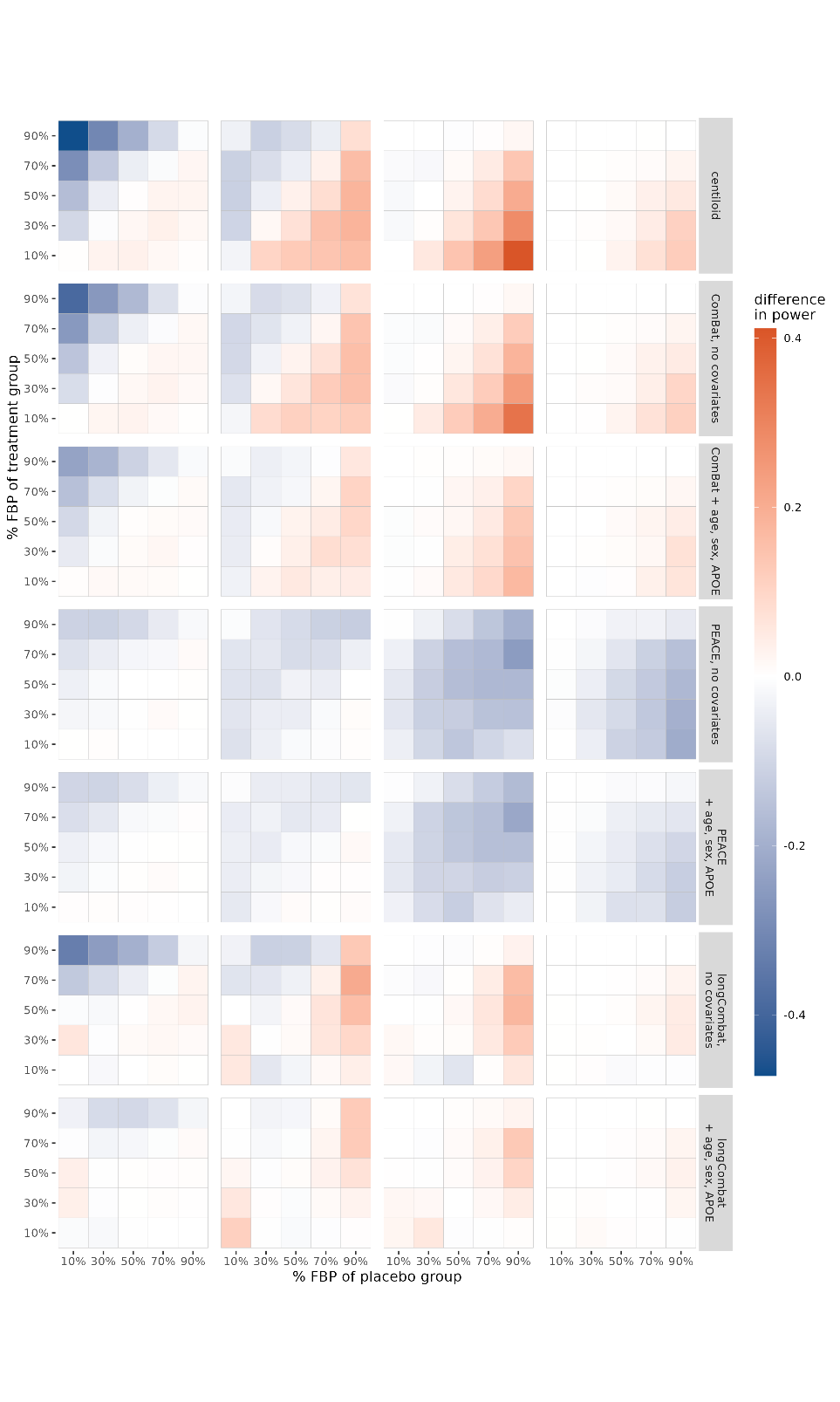

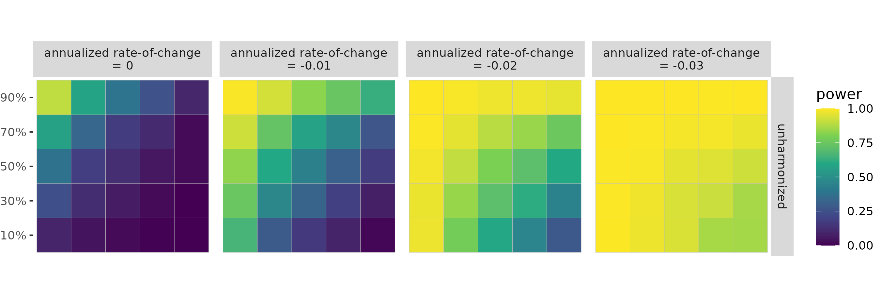

**Supplementary Fig. S7** Statistical power of detecting group differences in rate-of-change of the global summary SUVR between treatment and placebo groups, computed as the proportion of significant findings over 1000 iterations. Power is plotted for unharmonized SUVR, while difference in power relative to unharmonized is plotted for all harmonization methods. The true underlying rate-of-change is varied across columns. The proportion of FBP scans in the placebo and treatment groups are varied across the horizontal and vertical axes of each heatmap, respectively. Note that for annualized rate-of-change equal to zero, the proportion of significant findings corresponds to Type-I error rate. Heatmaps for unharmonized SUVR, Centiloid, ComBat without covariates, and ComBat with covariates are the same as Fig. 6 of the main text. Results for PEACE and longitudinal ComBat are also shown in this figure.

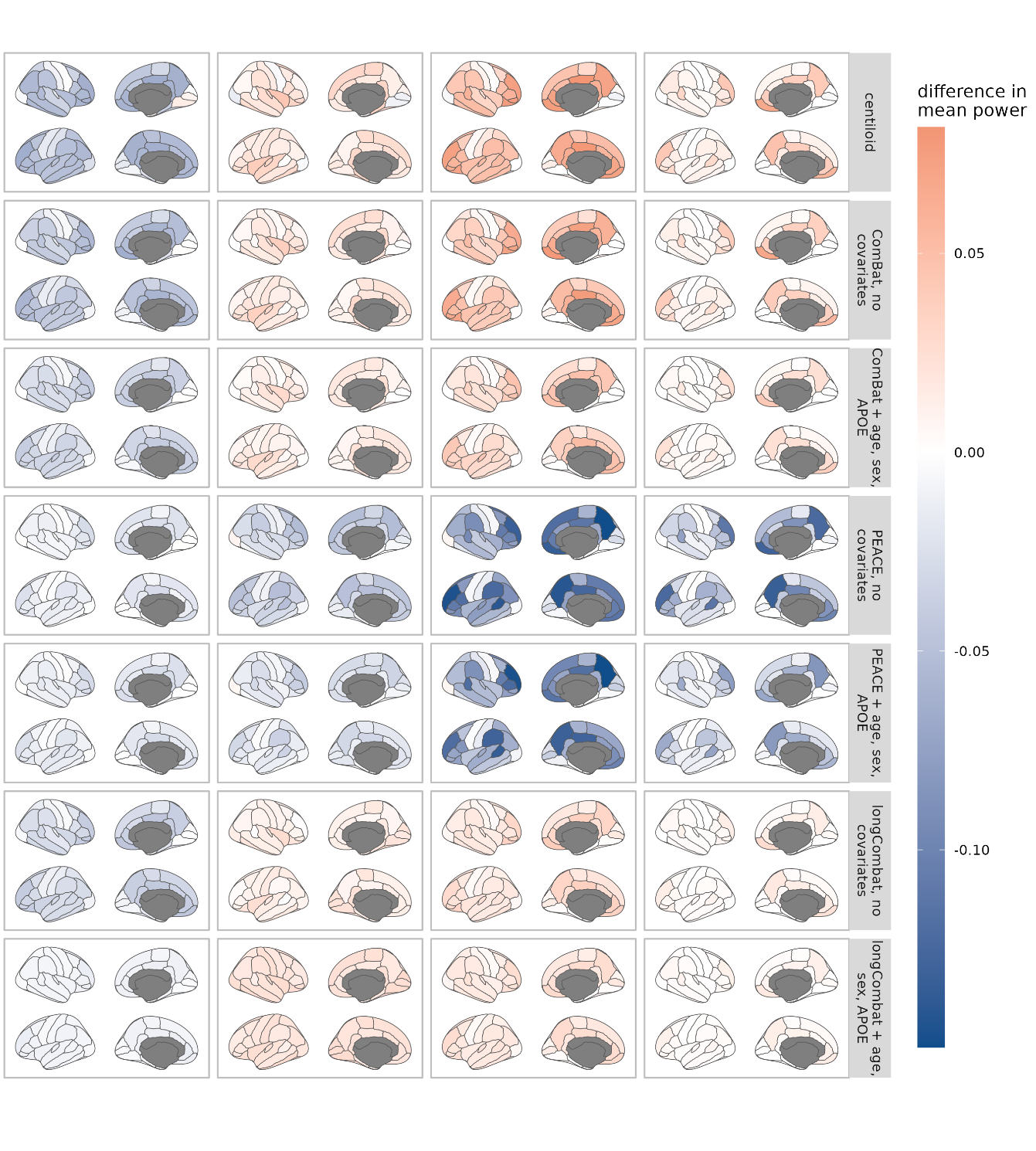

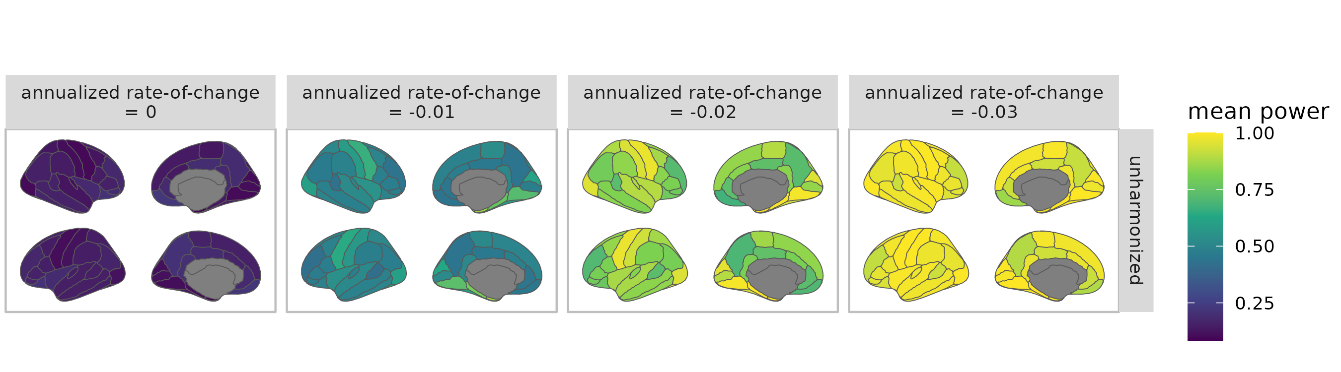

**Supplementary Fig. S8** Mean statistical power of detecting group differences in rate-of-change of cortical SUVRs between treatment and placebo groups, computed as the average across all permutations of tracer mixing proportions. Mean power is plotted for unharmonized SUVR, while difference in mean power relative to unharmonized is plotted for all harmonization methods. The true underlying rate-of-change is varied across columns. Note that for annualized rate-of-change equal to zero, the proportion of significant findings corresponds to Type-I error rate.

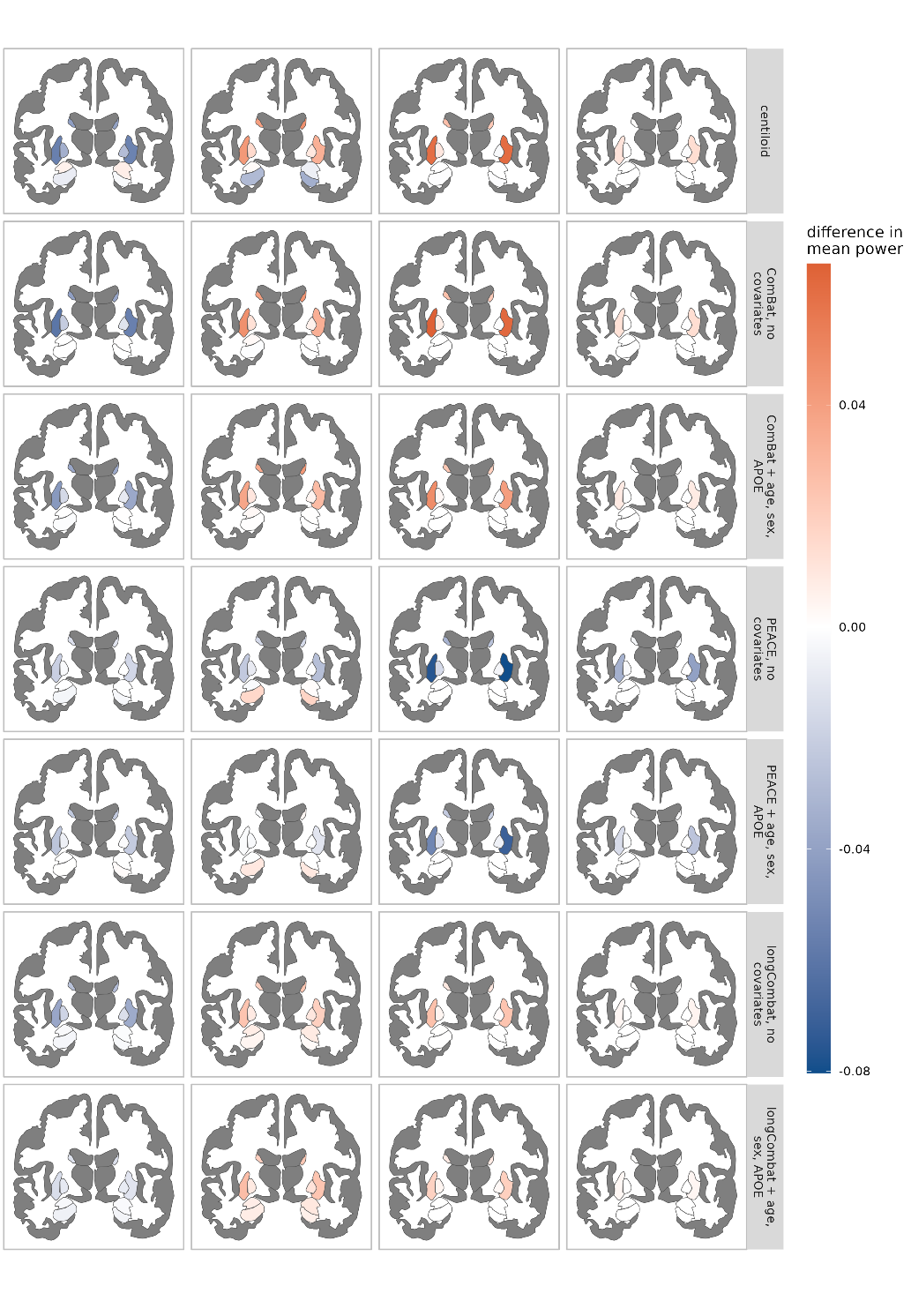

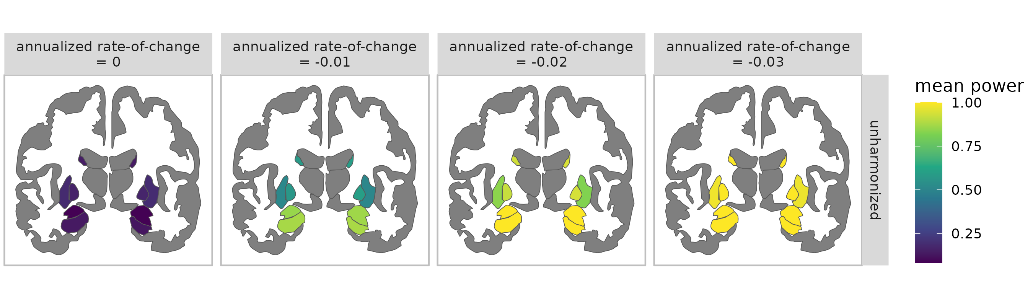

**Supplementary Fig. S9** Mean statistical power of detecting group differences in rate-of-change of subcortical SUVRs between treatment and placebo groups, computed as the average across all permutations of tracer mixing proportions. Mean power is plotted for unharmonized SUVR, while difference in mean power relative to unharmonized is plotted for all harmonization methods. The true underlying rate-of-change is varied across columns. Note that for annualized rate-of-change equal to zero, the proportion of significant findings corresponds to Type-I error rate.

**Supplementary Table S2** Mean statistical power of detecting significant rate-of-change differences between treatment and placebo groups in the simulation experiment for the global summary amyloid estimate. The mean and standard deviation were computed across all 25 combinations of tracer mixture proportions for treatment and placebo groups. The best performing harmonization methods are indicated in bold. Note that for the annualized rate-of-change of 0, lower power (i.e. Type-I error) corresponds to better performance.

|  | Mean power | | | |
| --- | --- | --- | --- | --- |
| Harmonization method | Annualized rate-of-change = 0 | Annualized rate-of-change = -0.01 | Annualized rate-of-change = -0.02 | Annualized rate-of-change = -0.03 |
| unharmonized | 0.161 ± 0.228 | 0.427 ± 0.314 | 0.735 ± 0.263 | 0.931 ± 0.095 |
| centiloid | **0.108 ± 0.105** | 0.512 ± 0.23 | **0.873 ± 0.115** | **0.986 ± 0.017** |
| ComBat, no covariates | 0.119 ± 0.129 | 0.511 ± 0.248 | 0.866 ± 0.13 | 0.985 ± 0.019 |
| ComBat + age, sex, APOE | 0.141 ± 0.171 | 0.505 ± 0.284 | 0.838 ± 0.176 | 0.976 ± 0.034 |
| PEACE, no covariates | 0.167 ± 0.192 | 0.448 ± 0.276 | 0.681 ± 0.23 | 0.887 ± 0.108 |
| PEACE + age, sex, APOE | 0.168 ± 0.192 | 0.472 ± 0.28 | 0.697 ± 0.22 | 0.921 ± 0.084 |
| longCombat, no covariates | 0.148 ± 0.148 | 0.51 ± 0.265 | 0.825 ± 0.188 | 0.97 ± 0.043 |
| longCombat + age, sex, APOE | 0.179 ± 0.213 | **0.518 ± 0.297** | 0.82 ± 0.2 | 0.969 ± 0.045 |

**Supplementary Table S3** Mean statistical power of detecting significant rate-of-change differences between treatment and placebo groups in the simulation experiment for the ROI amyloid measurements. To derive these statistics, the average power across all 25 combinations of tracer mixture proportions was computed for each ROI separately, then the mean and standard deviation of the ROI-specific average powers was computed across each of the ROI subgroups. The best performing harmonization methods are indicated in bold. Note that for the annualized rate-of-change of 0, lower power (i.e. Type-I error) corresponds to better performance.

|  | Mean power | | | |
| --- | --- | --- | --- | --- |
| Harmonization method | Summary cortical ROI | Other cortical ROI | Subcortical ROI | All ROI |
| **Annualized rate-of-change = 0** | | | | |
| unharmonized | 0.149 ± 0.032 | 0.114 ± 0.039 | 0.112 ± 0.034 | 0.12 ± 0.039 |
| centiloid | **0.106 ± 0.03** | **0.088 ± 0.017** | **0.099 ± 0.022** | **0.093 ± 0.022** |
| ComBat, no covariates | 0.113 ± 0.028 | 0.096 ± 0.02 | 0.102 ± 0.027 | 0.1 ± 0.023 |
| ComBat + age, sex, APOE | 0.132 ± 0.033 | 0.107 ± 0.026 | 0.108 ± 0.03 | 0.111 ± 0.029 |
| PEACE, no covariates | 0.158 ± 0.028 | 0.135 ± 0.027 | 0.131 ± 0.029 | 0.138 ± 0.029 |
| PEACE + age, sex, APOE | 0.158 ± 0.029 | 0.133 ± 0.026 | 0.128 ± 0.029 | 0.136 ± 0.029 |
| longCombat, no covariates | 0.142 ± 0.024 | 0.121 ± 0.021 | 0.12 ± 0.025 | 0.125 ± 0.023 |
| longCombat + age, sex, APOE | 0.167 ± 0.031 | 0.136 ± 0.03 | 0.131 ± 0.032 | 0.141 ± 0.032 |
| **Annualized rate-of-change = -0.01** | | | | |
| unharmonized | 0.419 ± 0.039 | 0.514 ± 0.122 | 0.582 ± 0.189 | 0.509 ± 0.134 |
| centiloid | 0.482 ± 0.044 | 0.558 ± 0.111 | 0.662 ± 0.154 | 0.562 ± 0.122 |
| ComBat, no covariates | 0.482 ± 0.042 | 0.561 ± 0.115 | **0.679 ± 0.159** | 0.567 ± 0.128 |
| ComBat + age, sex, APOE | 0.48 ± 0.039 | 0.558 ± 0.114 | 0.677 ± 0.161 | 0.565 ± 0.128 |
| PEACE, no covariates | 0.431 ± 0.041 | 0.522 ± 0.121 | 0.646 ± 0.179 | 0.528 ± 0.138 |
| PEACE + age, sex, APOE | 0.448 ± 0.039 | 0.532 ± 0.116 | 0.655 ± 0.17 | 0.539 ± 0.132 |
| longCombat, no covariates | 0.482 ± 0.036 | 0.56 ± 0.112 | 0.67 ± 0.16 | 0.565 ± 0.125 |
| longCombat + age, sex, APOE | **0.492 ± 0.034** | **0.566 ± 0.109** | 0.672 ± 0.159 | **0.572 ± 0.122** |
| **Annualized rate-of-change = -0.02** | | | | |
| unharmonized | 0.728 ± 0.096 | 0.846 ± 0.114 | 0.871 ± 0.136 | 0.83 ± 0.123 |
| centiloid | **0.841 ± 0.069** | **0.906 ± 0.067** | 0.938 ± 0.073 | **0.9 ± 0.073** |
| ComBat, no covariates | 0.837 ± 0.066 | 0.903 ± 0.067 | **0.946 ± 0.062** | 0.899 ± 0.073 |
| ComBat + age, sex, APOE | 0.814 ± 0.071 | 0.89 ± 0.076 | 0.938 ± 0.072 | 0.885 ± 0.082 |
| PEACE, no covariates | 0.674 ± 0.105 | 0.81 ± 0.128 | 0.886 ± 0.136 | 0.8 ± 0.14 |
| PEACE + age, sex, APOE | 0.685 ± 0.104 | 0.814 ± 0.124 | 0.893 ± 0.129 | 0.805 ± 0.136 |
| longCombat, no covariates | 0.804 ± 0.066 | 0.881 ± 0.079 | 0.93 ± 0.08 | 0.876 ± 0.085 |
| longCombat + age, sex, APOE | 0.8 ± 0.069 | 0.879 ± 0.081 | 0.928 ± 0.083 | 0.874 ± 0.087 |
| **Annualized rate-of-change = -0.03** | | | | |
| unharmonized | 0.911 ± 0.072 | 0.967 ± 0.045 | 0.969 ± 0.052 | 0.958 ± 0.055 |
| centiloid | **0.97 ± 0.026** | **0.988 ± 0.017** | 0.992 ± 0.015 | **0.986 ± 0.019** |
| ComBat, no covariates | **0.97 ± 0.025** | **0.988 ± 0.016** | **0.995 ± 0.01** | **0.986 ± 0.018** |
| ComBat + age, sex, APOE | 0.957 ± 0.034 | 0.983 ± 0.021 | 0.992 ± 0.013 | 0.981 ± 0.025 |
| PEACE, no covariates | 0.867 ± 0.088 | 0.946 ± 0.059 | 0.969 ± 0.054 | 0.937 ± 0.071 |
| PEACE + age, sex, APOE | 0.897 ± 0.071 | 0.958 ± 0.048 | 0.978 ± 0.039 | 0.951 ± 0.057 |
| longCombat, no covariates | 0.951 ± 0.036 | 0.98 ± 0.025 | 0.99 ± 0.019 | 0.976 ± 0.029 |
| longCombat + age, sex, APOE | 0.948 ± 0.039 | 0.979 ± 0.026 | 0.989 ± 0.02 | 0.975 ± 0.031 |
